# Understanding the Lived Experience of Chronic Pain: A Systematic Review and Synthesis of Qualitative Evidence Syntheses

**DOI:** 10.1101/2023.03.16.23287384

**Authors:** S van Rysewyk, R Blomkvist, R Crighton, F Hodson, D Roomes, E Shea, BH Smith, F Toye

## Abstract

**Background:** Although multiple measures of the causes and consequences of chronic non-cancer pain (CNCP) are available and can inform pain management, no quantitative summary of these measures can describe the meaning of pain for a patient. The lived experience of pain tends to be a blind spot in pain management. This study aimed to: (1) integrate qualitative research investigating the lived experience of a range of CNCP conditions; (2) establish common qualitative themes in CNCP experience; and (3) evaluate the relevance of our results through a survey questionnaire based on these themes, administered across the United Kingdom.

**Methods:** Six bibliographic databases were searched from inception to February 2021 to identify Qualitative Evidence Syntheses (QES) that investigated the lived experience of CNCP and its impact on everyday life and activities. Themes and trends were derived by thematic qualitative analysis in collaboration with two patient and public involvement representatives through two workshops. The output from these workshops helped inform the creation of twenty survey statements.

**Results:** The research team identified and screened 1,323 titles, and considered 86 abstracts, including 20 in the final review. Eight themes were developed from the study findings: (1) my pain gives rise to negative emotions; (2) changes to my life and to my self; (3) adapting to my new normal; (4) effects of my pain management strategies; (5) hiding and showing my pain; (6) medically explaining my pain; (7) relationships to those around me; and (8) working while in pain. Each theme gave rise to one or two survey questions. The survey was shared with members of the UK pain community over a two-week period in November 2021, and was completed by 1,219 people, largely confirming the above themes.

**Conclusion/implications:** This study provides a validated summary of the lived experience of CNCP. It highlights the adverse nature, complications, and consequences of living with CNCP in the UK, and the multiple shortcomings in the ways in which pain is addressed by others in the UK. Our findings are consistent with published meta-ethnographies on chronic non-malignant musculoskeletal pain, and chronic low-back pain. Despite the underrepresentation of qualitative research in the pain literature compared to quantitative approaches, for understanding the complexity of the lived experience of pain, qualitative research is an essential tool.

## Introduction

Pain is not simply an unpleasant immediate experience: over time, it can devastate a person’s health, quality of life, and it can end in suicide^1-6^. 10-year mortality is higher in people with severe chronic non-cancer pain (CNCP); death from heart or respiratory disease is twice the rate than people without severe CNCP^7^. Severe adverse effects are endured in families, employment, and societies of those with CNCP^8-10^. Chronic pain has been recognised internationally as a symptom or disease, with its inclusion in the 11^th^ Edition of the International Classification of Diseases (ICD-11)^11^. Chronic low-back pain was identified as a leading cause of disability worldwide, in both men and women^12^.

While medical scientists and academics work to solve the neurobiological questions of the origin, diagnosis, epidemiology and treatment of CNCP, such important work cannot capture the full meaning of a person’s lived experience of pain. The lived experience of CNCP remains a blind spot in pain management^13^. Patients often find medical explanations about pain unsatisfactory in relation to their lived experience of the pain^14^. Some patients find personal narratives and stories about pain, cautionary tales, and common-sense experience more meaningful and actionable than medical explanations^14^. This issue seems to be about meaning, where some people with pain do not find clinical discussions of pain meaningful, and clinicians typically are not looking for non-clinical, types of meaning. It has been suggested that one limitation for clinicians in treating pain and pain-related suffering effectively is an incomplete appreciation of the meaning of pain experience^15^.

Much of what is known today about the meaning and personal experiences of pain, has been achieved through qualitative research, which investigates important aspects of the pain experience that are relatively inaccessible to other approaches (e.g., quantitative approaches)^16,17^. Qualitative literature examining various aspects of living with an illness or chronic condition has increased dramatically in the last few years, with researchers finding that the number of relevant qualitative synthesis publications had doubled between 2005– 2008^18^.

Qualitative Evidence Syntheses (QES) bring together published qualitative research and develop ideas that cut across different contexts. There are several QES that explore aspects of the lived experience of chronic pain, and one study that provides a synthesis of QES^19-23^. These QES bring together more than 300 primary qualitative studies into a synthesised form. In this study, we planned a systematic search for existing QES that addressed the lived experience of CNCP, with the aim of synthesising the findings into themes that would underpin a survey questionnaire.

The aims of our study were to: (1) establish common qualitative themes in a range of CNCP conditions through a systematic review of published QES; and (2) to explore the relevance of our results by creating a survey questionnaire, based on these themes, across the United Kingdom.

## Methods

This study received approval by the University of Dundee School Research Ethics Committee (SMED REC Number 21/97). We used the methods of mega-ethnography developed by Toye et.al. to synthesise published Qualitative Evidence Syntheses (QES)^19-23^. Mega-ethnography follows the seven stages of meta-ethnography to synthesise research study findings^24^. There are no existing recommendations for reporting this type of review of reviews.

### Stage 1 (Getting started)

This stage incorporates the protocol development and project planning and oversight. The project was initiated by the organisation Pain UK, which is an umbrella organisation for UK-based charities that raise awareness and support people living with long-term pain. The project was overseen by a multidisciplinary steering committee of nine professionals working within or a relative field that is related in some way to those impacted by CNCP, including one clinical academic pain specialist (BS), one anthropologist/physiotherapist (FT), two nurses (FH, RC), one occupational medicine specialist (DR), one academic (SvR), and three patient and public involvement-engagement (PPIE) representatives (PG, ES, RC).

### Stage 2 (Deciding what is relevant)

We included all identified QES that explored the lived experience of CNCP. A systematic search of bibliographic databases was conducted from inception up until February 2021 using a combination of search terms developed to identify QES^19-23^ (Table 1). A single researcher screened titles, and the steering committee screened abstracts and agreed on full text inclusion.

**Table 1:** Search strategy.

|  |  |
| --- | --- |
| Sampling strategy | Comprehensive |
| Type of study | Qualitative Evidence Syntheses (QES) exploring the lived experience of CNCP |
| Approaches | Electronic database searches |
| Range of years (start date, end date) | From inception until February 2021 |
| Limits | Language: English |
| Inclusions and exclusions | Inclusion: Review of qualitative research; Lived experience of chronic pain (lasting more than 3 months); Explores the impact of pain on everyday life<br>Exclusion: Pain related to a specific time-limited health condition (e.g., cancer). Non-pain related experiences (such as stiffness, immobility) that could not be separated from the experience of pain; perspectives of family, carers, healthcare staff and employers where the data could not be separated from those of the person with pain. |
| Terms used | metasynthes* OR meta-synthes* OR "meta synthesis") OR (metasummar* OR |
|  | meta-summar* OR "meta summary") OR (metastud* OR meta-stud* OR "meta study") OR (metaethnog* OR meta-ethnog OR "meta ethnography") OR (metanarrative OR meta-narrative OR "meta narrative") OR "critical interpretive synthesis" OR (qualitative ADJ4 systematic*) OR (qualitative ADJ4 review) OR (qualitative ADJ4 syntheses*) combined with (exp PAIN or pain.ti, ab) |
| Electronic sources | APA, American Psychological Association; CINAHL, Cumulative Index to Nursing and Allied Health Literature; MEDLINE; PsycInfo |

**Table 2:** Qualitative Evidence Syntheses included in study.

| Author | Aim of QES | Search date | Number of studies | Analytic method | Analytic output |
| --- | --- | --- | --- | --- | --- |
| Bunzli et al. 2013<sup>[25]</sup> | To provide a richer understanding of what it is like to live with CLBP to encourage a move away from biomedical model | To October 2011 | 18 | Sandelowski and Barroso (coding, grouping and abstraction) | 11 categories and 3 themes |
| Chen et al. 2020<sup>[39]</sup> | To explore older adult's experiences of living with pain | To June 2019 | 11 | Noblit and Hare meta-synthesis | 25 metaphors and 4 themes |
| Crowe et al. 2017<sup>[38]</sup> | To synthesise descriptions of the experience of chronic pain across conditions | 2000-2015 | 41 | Thematic analysis and synthesis into meta-themes | 10 categories and 5 themes |
| Froud et al. 2014<sup>[26]</sup> | Synthesise the qualitative literature on the impact of low back pain on people's lives to suggest areas to explore in healthcare consultations | To July 2011 | 42 | Britten et.al (coding and meta-narrative) | 15 subthemes and 5 themes (plus second and third-order interpretations) |
| Grant et al. 2019<sup>[40]</sup> | Increase our understanding of the obstacles to returning to work for people with chronic pain and their employers | To April 2017 | 41 | Noblit and Hare meta-ethnography | 13 conceptual categories and 3 key categories |
| Khanom et al. 2020<sup>[35]</sup> | To identify knowledge gaps, and inform future research on pain flares by synthesising the literature on adolescents' experiences on fluctuating pain | To June 2018 | 32 | Thematic synthesis | 13 subthemes and 3 themes |
| MacNeela et al. 2015<sup>[27]</sup> | Synthesise research findings on the subjective meaning of CLBP | 1994-2011 | 28 | Noblit and Hare meta-ethnography | 13 subthemes and 4 themes |
| Maxwell et al. 2020<sup>[29]</sup> | Synthesise reviews exploring the experiences of individuals living with shoulder pain to enhance the understanding of these experiences, especially treatment-related experiences | To March 2020 | 26 | Noblit and Hare meta-ethnography | 3 themes |
| Nichols et al. 2017<sup>[30]</sup> | To systematically review the literature of the lived experience of chronic headache | 1988-2016 | 4 | Meta-ethnography | 3 themes |
| Shallcross et al. 2018<sup>[31]</sup> | Develop a broader understanding of women's experiences of vulvodynia by reviewing the qualitative literature | To January 2016 | 7 | Meta-ethnography | 11 sub-concepts and 4 key concepts |
| Sim et al. 2008<sup>[32]</sup> | Gain an interpretive understanding of the subjective impact of FMS by synthesising qualitative literature | To October 2006 | 23 | Meta-synthesis | 8 subthemes and 4 themes |
| Snelgrove et al. 2013<sup>[28]</sup> | To articulate the knowledge gained from reviewing qualitative studies of patients' experiences of CLBP to review authors recommendations | 2000-2012 | 28 | Meta-ethnography with thematic analysis and synthesis | 3 themes |
| Srisopa et al. 2020<sup>[33]</sup> | To develop a deeper understanding of the experience of living with PGP | 2000-2019 | 6 | Noblit and Hare meta-ethnography | 6 themes |
| Stewart et al. 2020<sup>[36]</sup> | To systematically review and thematically synthesise patient perspectives on gout flares to inform measures that represent flare burden | To 2019 | 16 | Meta-synthesis | 25 sub-themes and 4 themes |
| Souza et al. 2011<sup>[34]</sup> | To provide insight into CPP and form the basis for a biopsychosocial approach to the condition. | 1995-2010 | 7 | Meta-synthesis | 3 themes |
| Toye et al. 2013<sup>[23]</sup> | Synthesise reviews to improve understanding and thus best practice for people with MSK | To February 2012 | 60 | Noblit and Hare meta-ethnography | 6 conceptual categories and 1 theme |
| Toye et al. 2014<sup>[22]</sup> | Increase our understanding of patients' experiences of chronic pelvic pain | To March 2014 | 32 | Noblit and Hare meta-ethnography | 9 conceptual categories |
| Toye et al. 2016<sup>[21]</sup> | Increase our understanding of factors that limit people's ability to stay at work | To September 2021 | 19 | Noblit and Hare meta-ethnography | 5 conceptual categories |
| Vaismoradi et al. 2016<sup>[41]</sup> | To integrate international findings to enhance the understanding of experiences and perspectives on pain and pain management in nursing homes | "all years" | 6 | Noblit and Hare meta-ethnography | 6 sub-themes and 3 themes, 1 metaphor |
| Wride et al. 2018<sup>[37]</sup> | To identify and explore feeling and experiences of people living with knee pain in order to improve care | 2006-2016 | 9 | Contextualist approach and meta-aggregation | 10 categories and 2 synthesised themes |

**Table 3:** Themes identified in each Qualitative Evidence Synthesis.

| Author | My pain gives rise to negative emotions | Changes to my life and to my self | Adapting to my new normal | Effects of my pain management strategies | Hiding and showing my pain | Medically explaining my pain | Relationships to those around me | Working while in pain |
| --- | --- | --- | --- | --- | --- | --- | --- | --- |
| Bunzli et al. 2013<sup>[25]</sup> | X | X |  | X |  | X |  |  |
| Chen et al. 2020<sup>[39]</sup> | X |  | X | X | X |  |  |  |
| Crowe et al. 2017<sup>[38]</sup> | X | X | X |  | X | X |  |  |
| Froud et al. 2014<sup>[26]</sup> | X | X | X |  |  | X | X | X |
| Grant et al. 2019<sup>[40]</sup> |  | X |  |  | X |  | X | X |
| Khanom et al. 2020<sup>[35]</sup> | X | X | X |  | X |  | X |  |
| MacNeela et al. 2015<sup>[27]</sup> | X | X | X | X | X | X |  | X |
| Maxwell et al. 2020<sup>[29]</sup> | X | X |  | X |  | X | X |  |
| Nichols et al. (2017)<sup>[30]</sup> | X | X |  |  | X |  | X |  |
| Shallcross et al. (2018)<sup>[31]</sup> | X |  |  |  |  | X |  |  |
| Sim et al. (2008)<sup>[32]</sup> | X | X | X |  |  | X | X |  |
| Snelgrove et al. 2013<sup>[28]</sup> |  | X |  | X |  | X | X |  |
| Srisopa et al. 2020<sup>[33]</sup> | X | X | X |  |  | X |  |  |
| Stewart et al. 2020<sup>[36]</sup> | X | X | X |  |  | X | X |  |
| Souza et al. 2011<sup>[34]</sup> | X |  |  | X |  |  | X |  |
| Toye et al. 2013<sup>[23]</sup> | X | X |  |  | X | X | X |  |
| Toye et al. 2014<sup>[22]</sup> | X | X |  |  | X | X |  |  |
| Toye et al. 2016<sup>[21]</sup> |  |  |  |  |  |  |  | X |
| Vaismoradi et al. 2016<sup>[41]</sup> |  |  | X |  |  | X |  |  |
| Wride et al. 2018<sup>[37]</sup> | X | X |  |  |  | X |  |  |

### Stage 3: (Reading the studies)

Once the final studies had been selected and approved by the multidisciplinary steering committee, RB read each study and extracted the findings from the results sections into a spreadsheet. To facilitate communication of ideas, each finding was summarised and reworded as a first-person statement. This process usefully allowed the research team to focus on the essence of lived experience. This is a method previously used by Toye et.al.^19^ to facilitate understanding, empathy, and accessible language.

### Stage 4 and 5 (Determining how studies are related to each other and translating studies into each other)

These stages involved sorting review findings into themes through a process of constant comparison. The list of first-person statements developed in stage 3 was shared with PPIE representatives RC and PG who, together with RB, organised the statements into themes. RB, FT, and BS refined these themes through discussion.

### Stage 6 and 7 (Synthesising translations and expressing the synthesis)

These stages involved further refining the ideas through discussion and formatting these into a survey output.

### Developing the survey

Working with our PPIE members and advisory group, the team co-produced survey statements to reflect the final themes. One researcher (RB) drafted provisional statements and shared these with the full steering group through a draft survey. These were deliberated within the group through a series of consensus meetings to ensure that multiple views and disciplines were represented and included in the survey design. The survey questions were written in first-person formats that work well with a Likert scale (from strongly agree to strongly disagree).

The survey was created in Survey Monkey (Supplement One) and opened to the UK public between the 11^th^ and 28^th^ November 2021. The survey link was shared to Pain UK member charities via email and through their various social media networks. In November 2021, Pain UK had 40 member charities who collectively had an estimated 30,000 individual members signed up to their newsletters. The member charities ranged from large well-established organisations with thousands of members, to smaller and highly specialised charities focusing on specific conditions. Pain UK sent out the survey link to their 40 member charities, who had the option to forward the link to their individual members. They also shared the survey link through their social media profiles where it could be shared across the UK pain community.

## Results

### Qualitative Evidence Synthesis

RB identified and screened 1323 potential titles through the database search, of which 167 were uploaded to a citation manager software (EndNote) after initial title screening. After 81 duplicates were removed, 86 abstracts and 24 full texts were considered, with 20 studies included in the review (see Figure 1).

**Figure 1:**
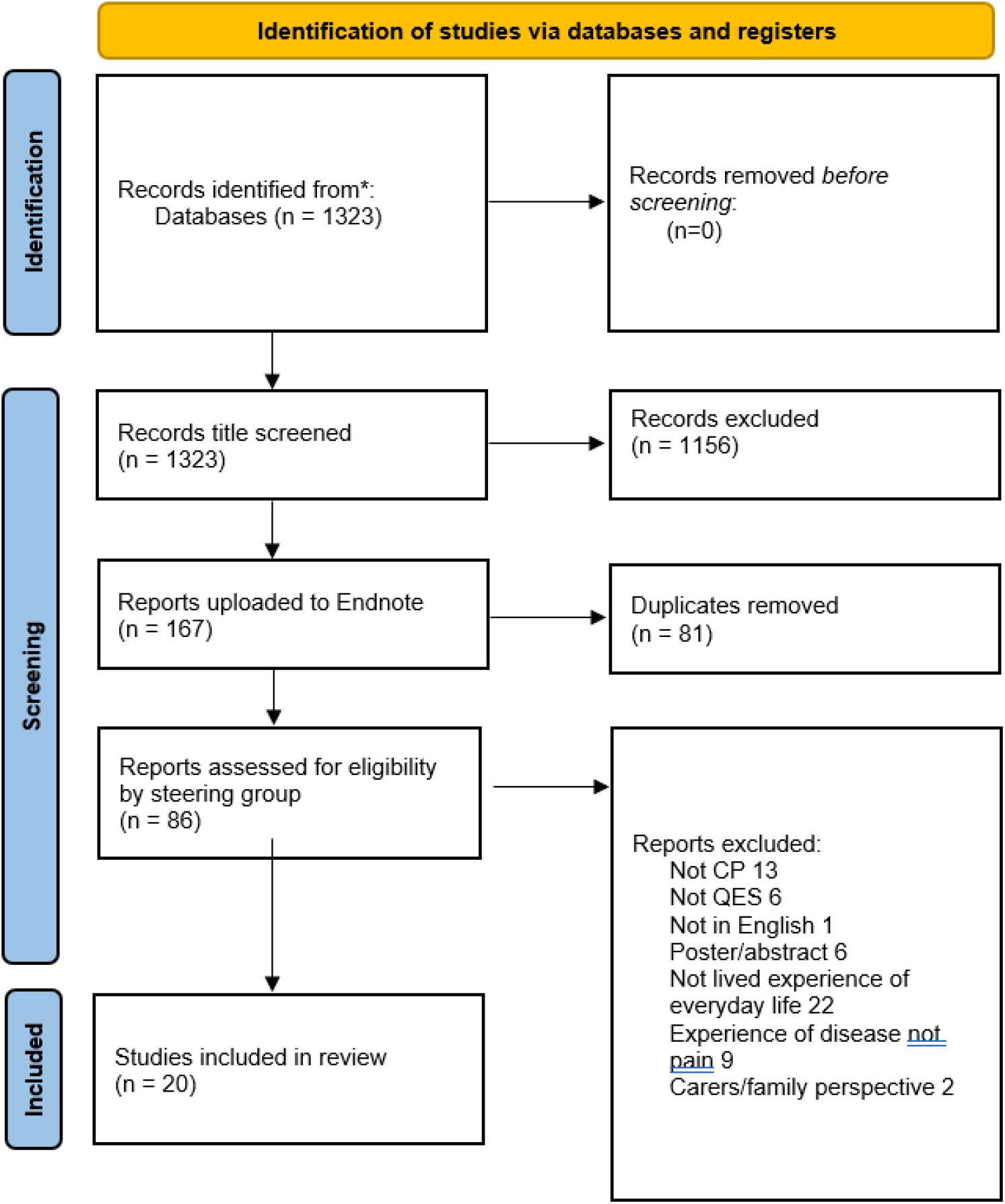
PRISMA Research Process Flow Diagram. *The 20 studies included in the review incorporated a range of pain conditions (low back pain, n = 4^[25-28]^; musculoskeletal pain, n = 2^[21,23]^; shoulder pain, n = 1^[29]^; headaches, n = 1^[30]^; vulvodynia, n = 1^[31]^; fibromyalgia, n = 1^[32]^; pelvic pain, n = 3^[22,33,34]^; pain flares, n = 2^[35,36]^; knee pain, n = 1^[37]^; and general chronic pain, n = 4^[38-41]^.

RB extracted 85 QES findings from 20 studies, which were organised into eight final themes and twenty survey statements. Table three shows which QES supported each theme. The themes are described below with two examples of findings from the primary studies supporting each theme. The examples show the first-person statement and are not narrative exemplars. Each theme is reported with its co-produced survey statement.

#### 1. My pain gives rise to negative emotions

This theme was supported by 15 out of 20 QES and describes the various aspects in which living with pain negatively affects mental health, and the deterioration of mental well-being because of living with chronic pain. The theme illustrates how the lived experience of pain can make a person’s life unpredictable, and induce feelings of anger, loss, or frustration.

> Crowe 2017^[38]^: I’m always attentive to my body to recognise early signs of pain: my pain remains unpredictable despite my efforts. I often feel overwhelmed and just surrender to my fate of having this illogical pain.
>
> Bunzli 2013^[25]^: My mood fluctuates between hope and despair, and I can see that this affects the people around me as well as my own mental health.

This theme was co-produced into the survey statement: “My mental health has deteriorated while living with pain”.

#### 2. Changes to my life and to my self

This theme was supported by 15 out of 20 QES and describes the impact on life and self: how living with pain changes the everyday activities and roles that can be performed and has a profound impact on expectations of the future, which together form a threat to one’s perceived self.

> MacNeela 2015^[27]^: Living with pain everyday has impacted my ability to do everyday tasks such as being a parent, caring for my garden, exercising my body, and driving my car. It feels like my life is not as full as it once was. Not being able to live in the way I did before makes me sad, and I worry about how this will be in the future if my condition doesn’t improve.
>
> Snelgrove 2013^[28]^: Living with pain has made me a different person and I mourn the “me” I used to be and the things I used to be able to do.

This theme was developed into two survey statements: “Since living with pain, I am no longer the person I used to be”, and “I am missing out on life because of my pain.”

#### 3. Adapting to my new normal

This theme was supported by 13 out of 20 QES and describes an attitude towards living with pain that accommodates pain without feeling that it limits life negatively. The theme also explores the social norms that might influence ideas about the level and type of pain that are considered “normal” regarding age and gender roles.

> Srisopa 2020^[33]^: I know I can never get my life back as it was before I developed pain. To move on, I try not to think about the pain. I have made changes in my lifestyle to accommodate the pain, I’ve lost weight, adjusted my activities, and sought medical help. I also try to think positively and accept the pain. Before I accepted the pain I struggled because I ignored the pain, avoided all movement, and didn’t understand where the pain came from. Now I have found a balance where I know I can’t do certain things and my overall pain has improved.
>
> Froud 2014^[26]^: After living with pain for some time, I understand and accept that I will not get the diagnosis I was hoping for, and that I need to adapt my lifestyle and accept that I live with pain.

This theme was developed into the survey statement: “Accepting that pain is part of my life would bring me some relief from the constant struggle to find a cure”.

#### 4. Negative effects of my pain management strategies

This theme was supported by 6 out of the 20 QES, and describes how some strategies to limit pain, such as avoidance of activities and self-medication, can have negative consequences on mental and social well-being, including isolation, depression, or alienation from others.

> Snelgrove 2013^[28]^: When I have bad pain I don’t think about long term strategies to end the pain, I just want the pain to end. Then I use medication and rest, sometimes even alcohol to feel better. I know these aren’t good long-term solutions, they make me isolated, depressed and it’s not a normal way to live a life, but in the moment, they are the only strategies that help.
>
> Bunzli 2013^[25]^: My condition is physical and I manage it by avoiding certain movements and activities, and by taking medication. Because I can no longer be who I want to be because of the pain, I am not as sociable as I used to be. This makes me isolated and depressed, but I prefer that to not feeling like I am myself around other people.

This theme was developed into the survey statement: “My pain management strategies improve my mental and social well-being”. After discussion with PPIE and advisors, the survey statement was co-produced as a mirror image of the theme finding, giving the respondents scope to rate it in either direction.

#### 5. Hiding and showing my pain

This theme was supported by 8 out of the 20 QES, and describes how living with pain can involve secrecy, shame, and hiding. The theme illustrates how, whilst hiding pain can lead to others not understanding or believing that pain exists, this strategy was sometimes chosen because exposing pain could lead to negative responses and judgement from others.

> Toye 2014^[22]^: I feel like I have to keep my condition secret and hidden because it’s not something you talk about. I would be embarrassed to mention it. But when I don’t talk about it, people don’t think it’s there, especially when I don’t have a diagnosis.
>
> Nichols 2017^[30]^: I struggle to plan ahead of time and it is therefore difficult to manage my relationships to my family, friends and wider community. Some friends I am closer to, now that I’ve shared my pain with them, but some relations have fallen by the wayside. Sometimes it’s helpful when people know about the pain, sometimes it makes me anxious. I make decisions about how much to disclose to people in different scenarios, sometimes I act normal to reduce other people being critical of me.

This theme was developed into the survey statement: “I sometimes hide my pain to avoid negative judgement from others”.

#### 6. Medically explaining my pain

This theme was supported by 14 out of 20 QES. It describes how the search for a diagnosis is associated with personal credibility, and reassurance that there is no malignant disease. It is underpinned by frustration towards the healthcare system and questioning about the legitimacy of pain as a biomedical condition.

> Bunzli 2013^[25]^: It is important for me to receive a medical explanation for my pain which I can understand and share and is acceptable to others. It is important to me that my family, employer, and welfare agent accept my condition as legitimate, and I will keep searching for a diagnosis that can satisfy this.
>
> Souza 2011^[34]^: I desperately want to know the cause of my pain so that I can know that it’s not dangerous and so that I can explain it to others so that they understand it is not in my head. No one can treat the pain if they don’t know what it is.

This theme was developed into the survey statement: “A formal diagnosis is important to me because it helps people around me believe and understand my pain is real”.

#### 7. Relationships to those around me

This theme was supported by 10 out of 20 QES. It describes the ways in which living with pain impacts social, professional, and familial relationships. The theme illustrates how these are especially influenced, either negatively or positively, by one’s ability to communicate effectively about pain.

> Grant 2019^[40]^: I don’t want to ask for help from my colleagues because it makes me feel inadequate and I don’t want to be a burden. I don’t trust people at my work not to judge me. When I don’t speak up about what I need it makes it harder to work, especially if my boss is unsympathetic.
>
> Khanom 2020^[35]^: Because no one can see my pain, I feel as though people around me think I am imagining it or that it’s not a legitimate issue to have. Sometimes I don’t talk about the pain because I want to avoid being judged.

This theme was developed into the survey statement: “I wish I could communicate more effectively with others about my pain”.

#### 8. Working while in pain

This theme was supported by 4 out of 20 QES. It conveys the challenge of asking for necessary support and modifications at work, largely due to fear of being viewed as workshy or dependent on others.

> Grant 2019^[40]^: I don’t want to ask for help from my colleagues because it makes me feel inadequate and I don’t want to be a burden. I don’t trust people at my work not to judge me. When I don’t speak up about what I need it makes it harder to work, especially if my boss is unsympathetic. Having poor relationships at work makes me less motivated to be in work.
>
> Toye 2016^[21]^: I used to feel really good about myself when I was at work, I was respected and valued. I’m very careful now to not be seen as a bad worker. To maintain my image, I struggle on despite the pain and sometimes rely on colleagues. I also take annual leave rather than sick days. I would rather quit my job than be seen as a bad worker.

This theme was developed into the survey statement: “My workplace provides me with appropriate support and modifications to enable me to perform at work despite my pain.” Again, after discussion with PPIE and advisors, the survey statement was co-produced as a mirror image of the theme finding, giving the respondents scope to rate it in either direction.

### The survey

1219 people completed the survey across the UK, using Survey Monkey (Table 4). The majority were from England (78.3%) and were female (89.5%). Most had a medical diagnosis to explain their pain (93%) and 50% of these had a diagnosis of fibromyalgia, with a large number indicating a diagnosis of endometriosis (22%). A relatively small percentage were diagnosed with arthritis or osteoarthritis (14%).

**Table 4:**
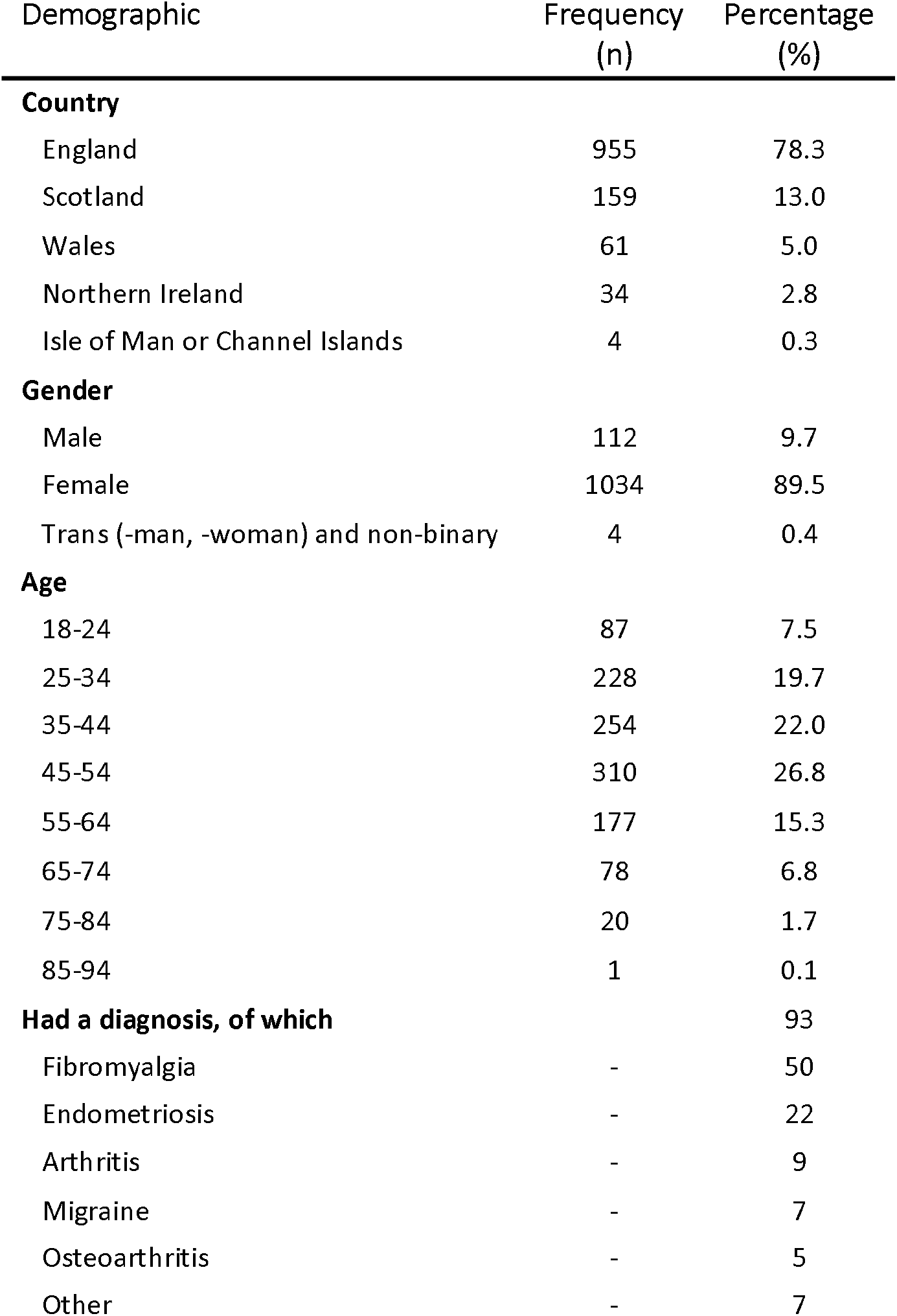
Demographics of survey respondents.

| Demographic | Frequency<br>(n) | Percentage<br>(%) |
| --- | --- | --- |
| **Country** |  |  |
| England | 955 | 78.3 |
| Scotland | 159 | 13.0 |
| Wales | 61 | 5.0 |
| Northern Ireland | 34 | 2.8 |
| Isle of Man or Channel Islands | 4 | 0.3 |
| **Gender** |  |  |
| Male | 112 | 9.7 |
| Female | 1034 | 89.5 |
| Trans (-man, -woman) and non-binary | 4 | 0.4 |
| **Age** |  |  |
| 18-24 | 87 | 7.5 |
| 25-34 | 228 | 19.7 |
| 35-44 | 254 | 22.0 |
| 45-54 | 310 | 26.8 |
| 55-64 | 177 | 15.3 |
| 65-74 | 78 | 6.8 |
| 75-84 | 20 | 1.7 |
| 85-94 | 1 | 0.1 |
| **Had a diagnosis, of which** |  | 93 |
| Fibromyalgia | - | 50 |
| Endometriosis | - | 22 |
| Arthritis | - | 9 |
| Migraine | - | 7 |
| Osteoarthritis | - | 5 |
| Other | - | 7 |

**Table 5:** Survey results.

| Theme | Question | Strongly agree (%) | Agree (%) | Neither agree nor disagree (%) | Disagree (%) | Strongly disagree (%) |
| --- | --- | --- | --- | --- | --- | --- |
| 1 | My mental health has deteriorated while living with pain. | 60% | 30% | 6% | 3% | 1% |
| 2a | I am missing out on my life because of my pain. | 59% | 32% | 6% | 3% | <0.5% |
| 2b | Since living with pain, I am no longer the person I used to be. | 59% | 27% | 9% | 4% | 1% |
| 3 | Accepting that pain is part of life would bring me some relief from the constant struggle to find a cure. | 25% | 31% | 25% | 15% | 5% |
| 4 | My pain management strategies improve my mental and social well-being. | 13% | 30% | 33% | 19% | 4% |
| 5 | I sometimes hide my pain to avoid negative judgement from others. | 63% | 29% | 4% | 3% | <0.5% |
| 6 | A medical diagnosis is important to me because it helps people around me believe and understand that my pain is real. | 65% | 23% | 7% | 3% | 1% |
| 7 | I wish I could communicate more effectively with others about my pain. | 54% | 30% | 12% | 4% | 1% |
| 8 | My pain affects my ability to perform at work because I do not receive the right support and modifications. | 33% | 23% | 31% | 10% | 3% |
| n/a | The COVID-19 pandemic has had a negative effect on my pain. | 33% | 20% | 29% | 14% | 4% |

The survey findings resonate with the themes from the mega-ethnography, and both highlight the adverse impact of CNCP on people’s lives in the UK. The vast majority of respondents either agreed, or strongly agreed, that their mental health had deteriorated (90%), that they were missing out on life (91%), or that they were no longer the person that they used to be (86%). The survey also highlighted the challenge of getting other people to understand, and believe in, your pain: 92% hid pain to avoid judgement and 88% felt that a diagnosis would help people to believe and understand pain, and 84% wanted to be able to communicate more effectively with others about their pain. Fewer than half of the respondents (43%) felt that their pain management strategies improved their mental and social wellbeing and 23% felt that these strategies did not help. See Supplement One for the full survey results.

## Discussion

### Summary of study

This study identified QES that systematically explored and explained the experience of living with chronic non-cancer pain in the UK. We identified 85 separate themes described in the reviewed QES studies, and grouped them into 8 qualitative themes. We tested these themes in a survey using a large group of people living with CNCP in the UK, and found strong, or very strong, agreement, with each of them. This study therefore provides a comprehensive, validated summary of the experience of living with CNCP, particularly in the UK.

Despite the increasing number of qualitative studies on pain, the lived experience of pain remains a blind spot in the field because qualitative inquiry continues to be underrepresented in the pain literature in relation to quantitative approaches, which are underpinned by the predominant biomedical approach to pain management. In understanding the complexity of the lived experience of pain, qualitative research is an indispensable tool^16,17^.

### Interpretation

Our findings reveal the adverse nature, complications, and consequences of living with chronic non-cancer pain among people in the UK. People living with CNCP often struggle to make sense of the consequences of pain, which disrupts the sense of who they are and what they can do (Themes 1-3). The challenge in effectively communicating their pain experience to others, and the consequent misunderstanding of other people, including healthcare professionals, and work colleagues, further exacerbates the adversity of pain (Themes 7, 8).

For many people with chronic pain, a medical diagnosis is important because it “helps people around me believe and understand that my pain is real” (Theme 6). This emphasises the importance of early diagnosis of chronic pain when possible, and of the relative contribution of physical, psychological, or environmental factors, to ongoing pain management^42,43^.

The findings presented in this paper point to shortcomings in the ways in which pain is addressed by others in the UK (Themes 4-8). People with pain, their family, or carers, should have the knowledge and confidence to search for advice, education, or treatment to better understand and manage their pain. People who can make sense of their pain and can integrate this new reality into their self-concept are better able to move forward in life (Themes 1-3)^44^.

For medical, nursing, and allied health professionals, education and training in pain management provides the knowledge and resources to deliver best-practice, evidence-based care of pain. Education in the biopsychosocial sciences underpinning chronic pain might give health professionals an accurate conceptualisation of pain and readiness to hear patient narratives of chronic pain, and to incorporate this knowledge into treatment approaches (Themes 4, 6, 7)^45,46^.

To facilitate and encourage engagement with this topic by medical or health care practitioners, it would be beneficial to improve training opportunities for those interested in using qualitative methods (Theme 4).

### Comparison with previous literature

The present study adds to the existing literature by synthesising the findings of studies that explore the lived experience of CNCP in everyday life from a variety of contexts, including a range of study samples, geographical areas, ages, and pain conditions. Our findings are broadly consistent with results in published meta-ethnographies on chronic non-malignant musculoskeletal pain^23,47^, and chronic low-back pain^25-27^. For example, Toye et al.^23,47^ identified chronic pain as an “adversarial struggle”, consisting of five qualitative themes. People with pain struggle to affirm themselves (Themes 1, 2), adapt to living with pain (Theme 3), find a suitable explanation for their pain (Theme 6), negotiate the healthcare system (Theme 7), and be viewed by observers as legitimate (Themes 6, 7). Beyond the meta/mega-ethnography, our results align with published data in other qualitative systematic or meta-analyses on chronic pain^38,48,49^. The main innovation of our study is the development of a survey to evaluate our findings in a UK population.

## Strengths and limitations

Our study uses a systematic research method that has been used to synthesise QES for rheumatoid arthritis and chronic pain^24,50,51^, yet there are no reporting guidelines for reviews of QES. Our findings, although extensive, are not necessarily all-inclusive: this is not incompatible with interpretive methodologies that aim to develop ideas. The inclusion of PPIE in our research design, conduct, and analysis was a key strength, ensuring that stakeholder views and perspectives were represented^52,53^, and played a key role in analysis and design of survey questions. A strength of our design was the validation of our findings through the creation and use of a survey in the pain community, which confirmed the clinical relevance of our findings, and the importance of qualitative research on pain^16,17^. Most survey respondents had a diagnosed chronic pain condition, including 22% with diagnosed endometriosis, which is sometimes described as the “missed disease” due to its unclear aetiology, or inconsistencies in diagnosis and management^54^.

Potential limitations include evaluating only English language studies. However, only one non-English language study was excluded, and the six databases consulted are the most relevant. Furthermore, the large number of duplicates identified in our search supports the comprehensiveness of our strategy. A further possible limitation is the lack of representativeness of the sample participating in the survey, compared with the UK population living with pain, for example 22% with endometriosis. As the survey was sent to particular chronic pain organisations, some CNCP conditions (such as endometriosis and fibromyalgia) may have been overrepresented, and others (such as joint pain or low-back pain) were underrepresented, the reasons for which were out of our control. However, the strength of our findings is that a wide range of conditions and demographics are represented in the survey, adding to our knowledge of pain, though cannot be taken to represent the general population. Other research has found that the number of sites of chronic pain may be more important than the actual site(s), or diagnosis of chronic pain, in determining its impact on lived experience^55^. Further research to explore the impact of pain in specific pain conditions and in specific socio-economic contexts would add to our knowledge of pain.

## Conclusion

Qualitative research is indispensable in understanding the lived experience of chronic pain. Our systematic review of QES and patient survey describes the adverse experience, complications, and consequences of chronic non-cancer pain. People with pain, their significant others, or carers, should have the knowledge and assurance to seek advice, education, or treatment to understand and manage their pain. Education and ongoing training in the biopsychosocial model of CNCP should provide health professionals with an accurate understanding of pain and preparedness to understand and support the patient’s account of their lived pain. Recognition and personalised clinical management for people living with CNCP should be pursued as a health priority in the UK.

## Supporting information

Supplemental Survey

## Data Availability

All data produced in the present study are available upon reasonable request to the authors.

## Supplementary files

Full survey (Supplement 1)

## Acknowledgements

The authors wish to acknowledge Antony Chuter and Peter Gregory for their time developing and reviewing the manuscript.

## References

1. Maher C, Underwood M, Buchbinder R. Non-specific low back pain. Lancet 2017; 389: 736–47.

2. Chenot JF, Greitemann B, Kladny B, et al. Non-specific low back pain. Dtsch Arztebl Int 2017; 114: 883–90.

3. Elliott AM, Smith BH, Penny KI, et al. The epidemiology of chronic pain in the community. Lancet 1999; 354: 1248–1252.

4. Smith BH, Elliott AM, Chambers WA, et al. The impact of chronic pain in the community. Fam Pract 2001; 18: 292–299.

5. Racine M. Chronic pain and suicide risk: A comprehensive review. Prog Neuropsychopharmacol Biol Psych. 2018; 20: 269–80.

6. Pergolizzi Jr JV, Magnusson P, LeQuang JA, et al. Suicide, opioids, chronic pain, and mental health disorders: a narrative review. Addict Sub Abuse 2022; 1: 15–9.

7. Torrance, N, Elliott AM, Lee AJ, et al. Severe chronic pain is associated with increased 10 year mortality. A Cohort record linkage study. Euro J Pain 2010; 14: 380–386.

8. Ojeda B, Salazar A, Dueñas M, et al. The impact of chronic pain: the perspective of patients, relatives, and caregivers. Fam Syst Health 2014; 32: 399.

9. Patel AS, Farquharson R, Carroll D, et al. The impact and burden of chronic pain in the workplace: a qualitative systematic review. Pain Pract 2012; 12: 578–89.

10. Prego-Domínguez J, Khazaeipour Z, Mallah N, et al. Socioeconomic status and occurrence of chronic pain: a meta-analysis. Rheumatol 2021; 60: 1091–105.

11. Treede RD, Rief W, Barke A, et al. Chronic pain as a symptom or a disease: the IASP Classification of Chronic Pain for the International Classification of Diseases (ICD-11). Pain 2019; 160: 19–27.

12. James SL, Abate D, Abate KH, et al. Global, regional, and national incidence, prevalence, and years lived with disability for 354 diseases and injuries for 195 countries and territories, 1990–2017: a systematic analysis for the Global Burden of Disease Study 2017. Lancet 2018; 392: 1789–858.

13. Price DD, Barrell JJ. Inner experience and neuroscience: merging both perspectives. MIT Press; 2012.

14. Frank AW. Asking the right question about pain: Narrative and phronesis. Lit Med 2004; 23: 209–25.

15. van Rysewyk S, Galbraith M, Quintner J, et al. Do we mean to ignore meaning in pain? Pain Med 2021; 22: 1021–3.

16. Tutelman PR, Webster F. Qualitative research and pain: Current controversies and future directions. Can J Pain 2020; 4: 1–5.

17. Osborn M, Rodham K. Insights into pain: a review of qualitative research. Rev Pain 2010; 4: 2–7.

18. Hannes K, Macaitis K. A move to more systematic and transparent approaches in qualitative evidence synthesis: update on a review of published papers. Qual Res 2012; 12: 40–42.

19. Toye F, Belton J, Hannink E, et al. A healing journey with chronic pain: a meta-ethnography synthesizing 195 qualitative studies. Pain Med 2021; 22: 1333–44.

20. Toye F, Seers K, Barker K, et al. A mega-ethnography of eleven systematic reviews of qualitative research exploring the experience of living with pain. Brit J Pain 2017; 11: 9–10.

21. Toye F, Seers K, Allcock N, et al. A synthesis of qualitative research exploring the barriers to staying in work with chronic musculoskeletal pain. Disabil Rehab 2016; 38: 566–572.

22. Toye F, Seers K, Barker K. A meta-ethnography of patients’ experiences of chronic pelvic pain: struggling to construct chronic pelvic pain as ‘real’. J Adv Nurs 2014; 70: 2713–2727.

23. Toye F, Seers K, Allcock N, et al. Patients’ experiences of chronic non-malignant musculoskeletal pain: a qualitative systematic review. Brit J Gen Pract 2013; 63: e829–41.

24. Noblit, GW, Hare RD. Meta-Ethnography. Qualitative Research Methods. Thousand Oaks, CA: SAGE Publications, Inc., 1988.

25. Bunzli S, Watkins R, Smith A, et al. Lives on Hold: A Qualitative Synthesis Exploring the Experience of Chronic Low-back Pain. Clin J Pain 2013; 29: 907–916.

26. Froud R, Patterson S Eldridge, et al. A systematic review and meta-synthesis of the impact of low back pain on people’s lives. BMC Musculoskel Disord 2014; 15: 1–4.

27. MacNeela P, Doyle C, O’Gorman D, et al. Experiences of chronic low back pain: a meta-ethnography of qualitative research. Health Psych Rev 2015; 9: 63–82.

28. Snelgrove S, Liossi C. Living with chronic low back pain: a metasynthesis of qualitative research. Chron Ill 2013; 9: 283–301.

29. Maxwell CK, Robinson K, McCreesh K. Understanding Shoulder Pain: A Qualitative Evidence Synthesis Exploring the Patient Experience. Phys Thera 2020; 28: 229.

30. Nichols VP, Ellard DR, Griffiths FE. The lived experience of chronic headache: a systematic review and synthesis of the qualitative literature. BMJ Open 2017; 7: e019929.

31. Shallcross R, Dickson JM, Nunns D, et al. Women’s Subjective Experiences of Living with Vulvodynia: A Systematic Review and Meta-Ethnography. Arch Sex Behav 2018; 47: 577–595.

32. Sim J, S Madden. Illness experience in fibromyalgia syndrome: a metasynthesis of qualitative studies. Soc Sci Med 2008; 67: 57–67.

33. Srisopa P, Lucas R. Women’s Experience of Pelvic Girdle Pain After Childbirth: A Meta-Synthesis. J Midwif Women Health 2020; 14: 14.

34. Souza PP, Salata Romão A, Rosa-e-Silva JC, et al. Qualitative research as the basis for a biopsychosocial approach to women with chronic pelvic pain. J Psychosom Obstet Gynaecol 2011; 32: 165–72.

35. Khanom S, McDonagh JE, Briggs M, et al. Adolescents’ experiences of fluctuating pain in musculoskeletal disorders: a qualitative systematic review and thematic synthesis. BMC Musculoskel Disord 2020; 21: 1–16.

36. Stewart, S, Guillen AG, Taylor WJ, et al. The experience of a gout flare: a meta-synthesis of qualitative studies. Sem Arthr Rheum 2020; 50: 805–811.

37. Wride JM, Bannigan K. ‘If you can’t help me, so help me God I will cut it off myself…’ The experience of living with knee pain: a qualitative meta-synthesis. Physiothera 2018; 104: 299–310.

38. Crowe M, Whitehead L, Seaton P, et al. Qualitative meta-synthesis: the experience of chronic pain across conditions. J Adv Nurs 2017; 73: 1004–1016.

39. Chen J, Hu F, Yang BX, et al. Experience of living with pain among older adults with arthritis: A systematic review and meta-synthesis. Internat J Nurs Stud 2020; 111: 103756.

40. Grant, M, Joanne O, Froud R, et al. The work of return to work. Challenges of returning to work when you have chronic pain: a meta-ethnography. BMJ Open 2019; 9: e025743.

41. Vaismoradi, M, Skär L, Söderberg S, et al. Normalizing suffering: A meta-synthesis of experiences of and perspectives on pain and pain management in nursing homes. Internat J Qualit Stud Health Well-Being 2016; 11: 31203.

42. Cush JJ. Rheumatoid arthritis: early diagnosis and treatment. Med Clin 2021; 105: 355–65.

43. McHugh G, Thomas G. Living with chronic pain: the patient’s perspective. Nurs Stand 2001; 15: 33–7.

44. Lennox Thompson B, Gage J, Kirk R. Living well with chronic pain: a classical grounded theory. Disabil Rehab 2020; 42: 1141–52.

45. Ng W, Slater H, Starcevich C, et al. Barriers and enablers influencing healthcare professionals’ adoption of a biopsychosocial approach to musculoskeletal pain: a systematic review and qualitative evidence synthesis. Pain 2021; 162: 2154–85.

46. Holopainen R, Simpson P, Piirainen A, et al. Physiotherapists’ perceptions of learning and implementing a biopsychosocial intervention to treat musculoskeletal pain conditions: a systematic review and metasynthesis of qualitative studies. Pain 2020; 161: 1150–68.

47. Toye F, Seers K, Allcock N, et al. A meta-ethnography of patients’ experience of chronic non-malignant musculoskeletal pain. Health Serv Deliv Res 2013; 1: S259–60.

48. Falsiroli Maistrello L, Zanconato L, Palese A, et al. Perceptions and Experiences of Individuals With Neck Pain: A Systematic Critical Review of Qualitative Studies With Meta-Summary and Meta-Synthesis. Physical thera 2022; 102: pzac080.

49. Devan H, Hale L, Hempel D, et al. What works and does not work in a self-management intervention for people with chronic pain? Qualitative systematic review and meta-synthesis. Physical thera 2018; 98: 381–97.

50. Toye F, Seers K, Hannink E, et al. A mega-ethnography of eleven qualitative evidence syntheses exploring the experience of living with chronic non-malignant pain. BMC Med Res Methodol 2017; 17: 1.

51. Toye F, Seers K, Barker K. Living life precariously with rheumatoid arthritis - a mega-ethnography of nine qualitative evidence syntheses. BMC Rheumatol 2019; 3: 1–3.

52. Babatunde OO, Dawson S, Brammar J, et al. Patient and public involvement in implementation of evidence-based guidance for musculoskeletal conditions: a scoping review of current advances and gaps. BMC Rheumatol 2022; 6: 1–23.

53. Holzer KJ, Veasley C, Kerns RD, et al. Partnering with patients in clinical trials of pain treatments: a narrative review. Pain 2022; 28: 10–97.

54. Hudson N. The missed disease? Endometriosis as an example of ‘undone science’. Reprod Biomed Soc Online 2022; 14: 20–7.

55. Smith BH, Elliott AM, Hannaford PC. Is chronic pain a distinct diagnosis in primary care? Evidence arising from the Royal College of General Practitioners’ Oral Contraception study. Fam Pract 2004; 21: 66–74.

