## Supplemental Survey for "Understanding the Lived Experience of Chronic Pain: A Systematic Review and Synthesis of Qualitative Evidence Syntheses"

### Slide 1
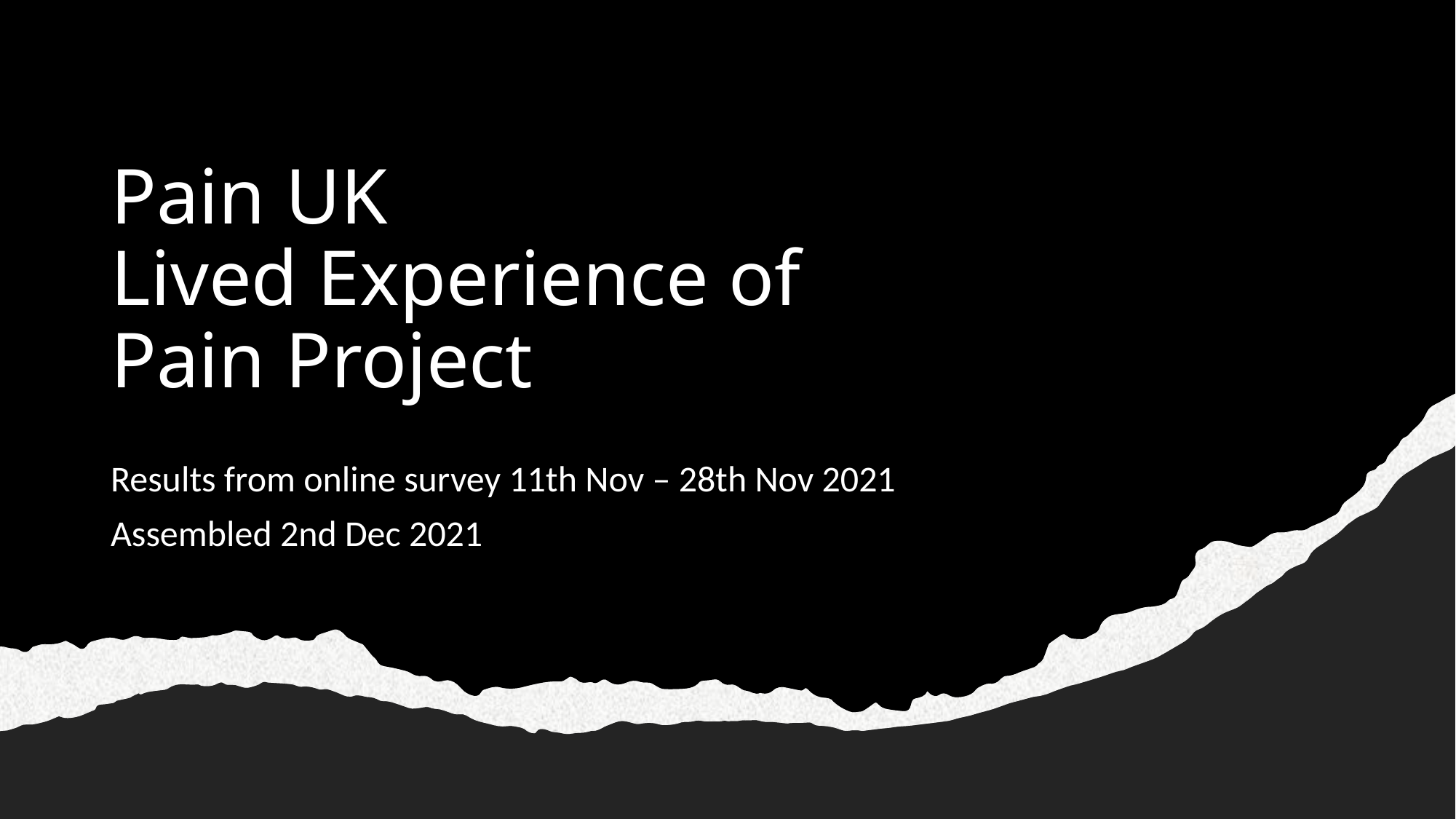

#### Pain UK Lived Experience of Pain Project
Results from online survey 11th Nov – 28th Nov 2021
Assembled 2nd Dec 2021

### Slide 2
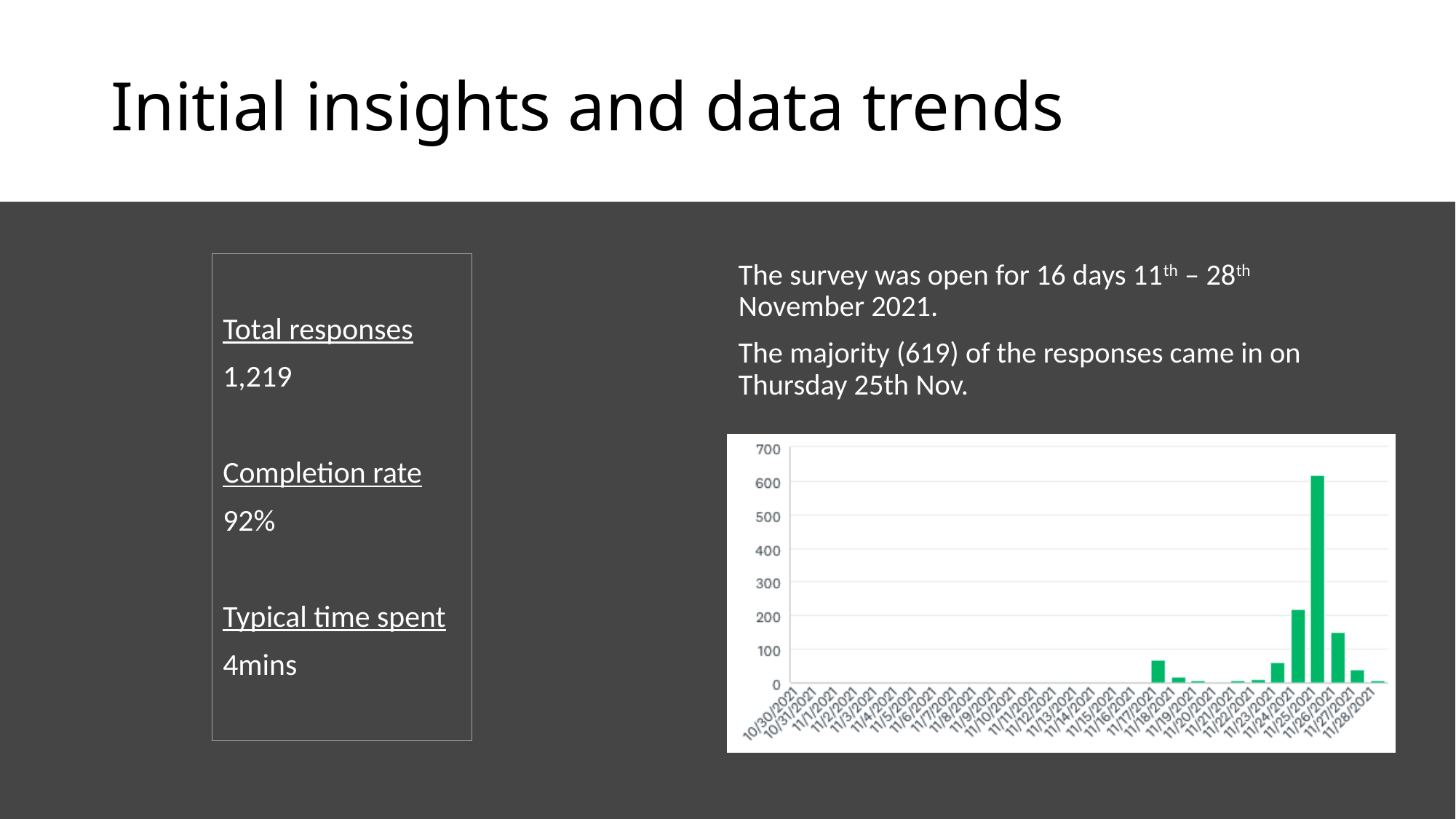

#### Initial insights and data trends
Total responses
1,219
Completion rate
92%
Typical time spent
4mins
The survey was open for 16 days 11^th^ – 28^th^ November 2021.
The majority (619) of the responses came in on Thursday 25th Nov.

### Slide 3
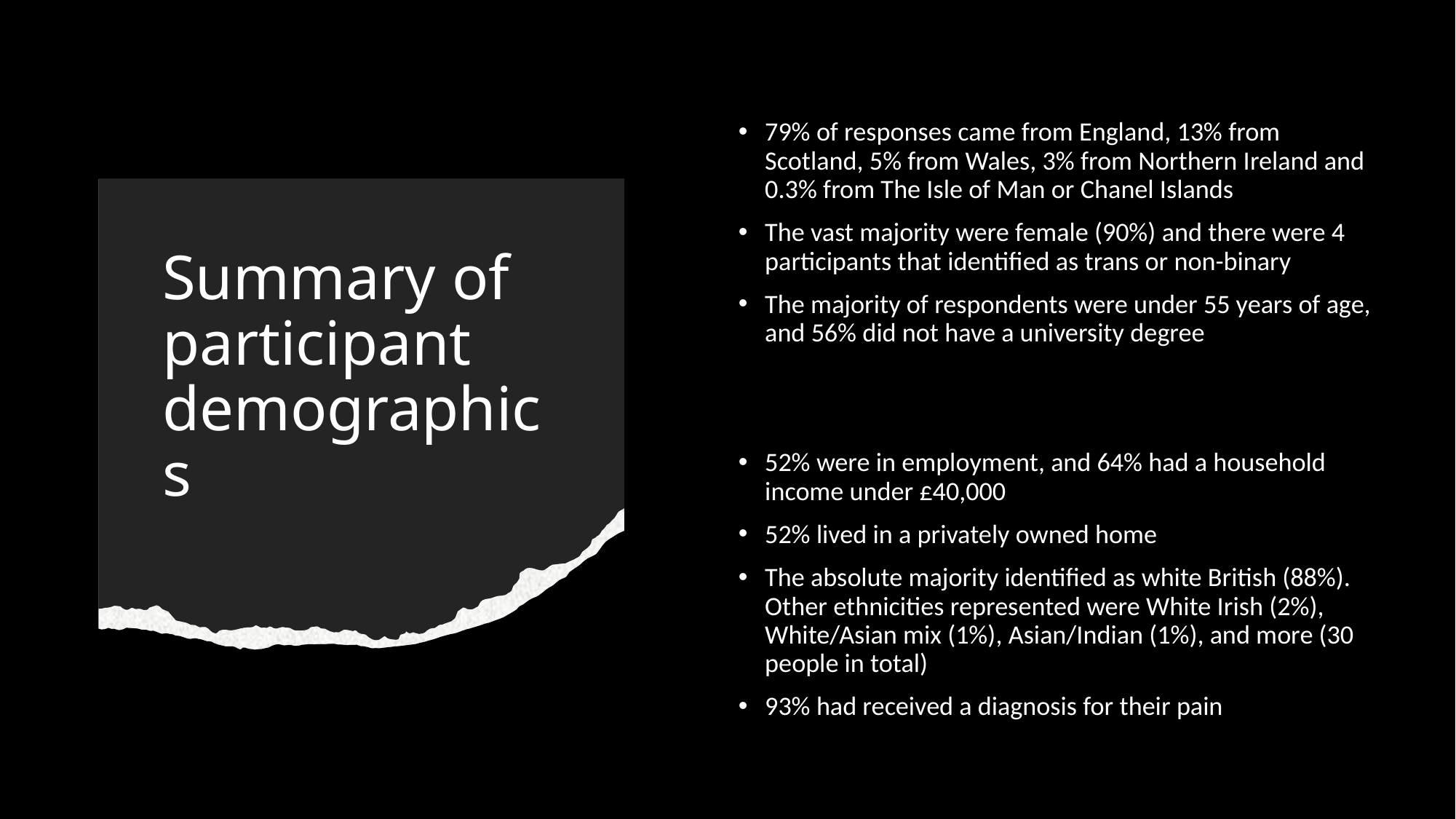

79% of responses came from England, 13% from Scotland, 5% from Wales, 3% from Northern Ireland and 0.3% from The Isle of Man or Chanel Islands
The vast majority were female (90%) and there were 4 participants that identified as trans or non-binary
The majority of respondents were under 55 years of age, and 56% did not have a university degree
52% were in employment, and 64% had a household income under £40,000
52% lived in a privately owned home
The absolute majority identified as white British (88%). Other ethnicities represented were White Irish (2%), White/Asian mix (1%), Asian/Indian (1%), and more (30 people in total)
93% had received a diagnosis for their pain
#### Summary of participant demographics

### Slide 4
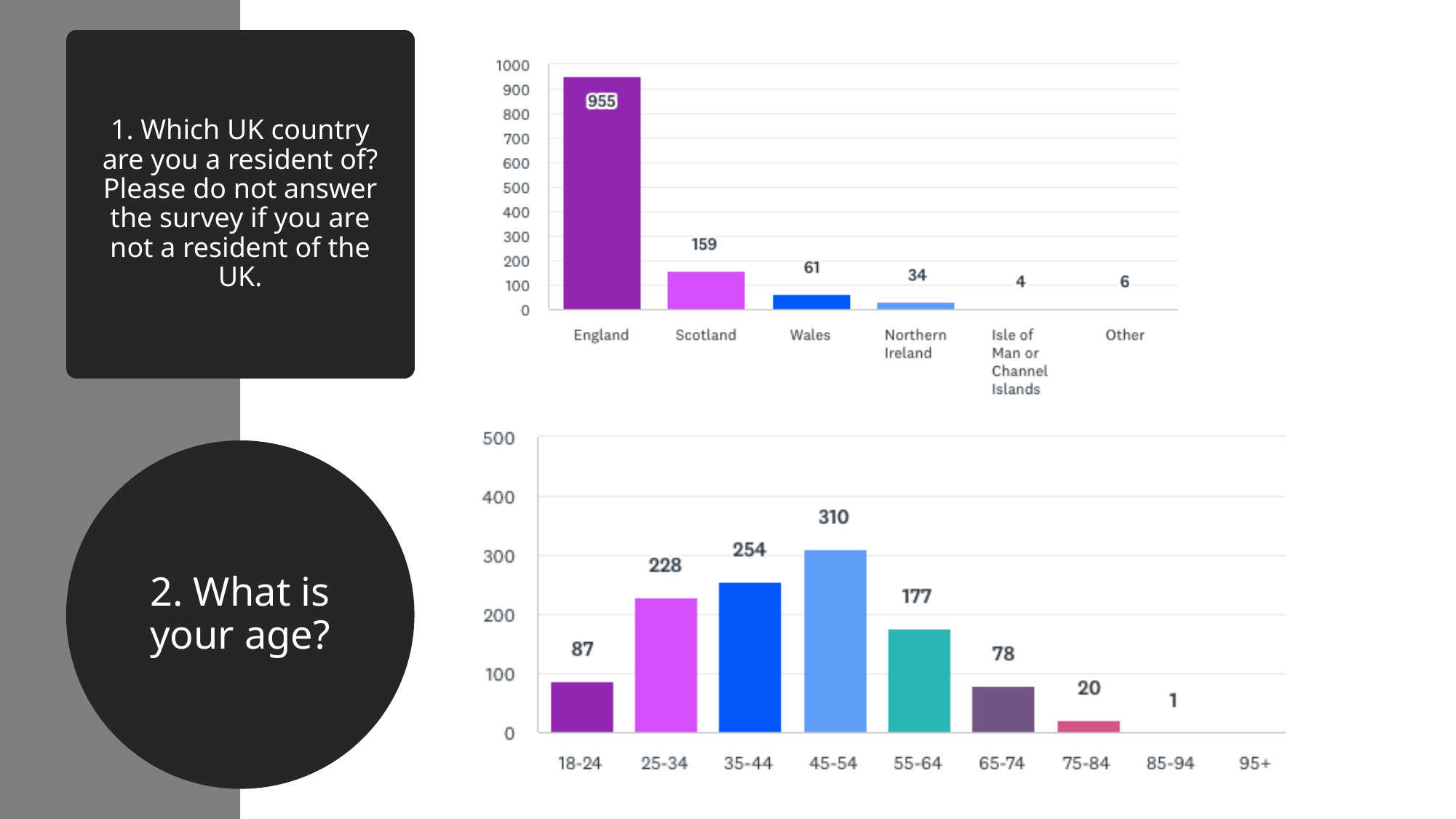

#### 1. Which UK country are you a resident of? Please do not answer the survey if you are not a resident of the UK.
2. What is your age?

### Slide 5
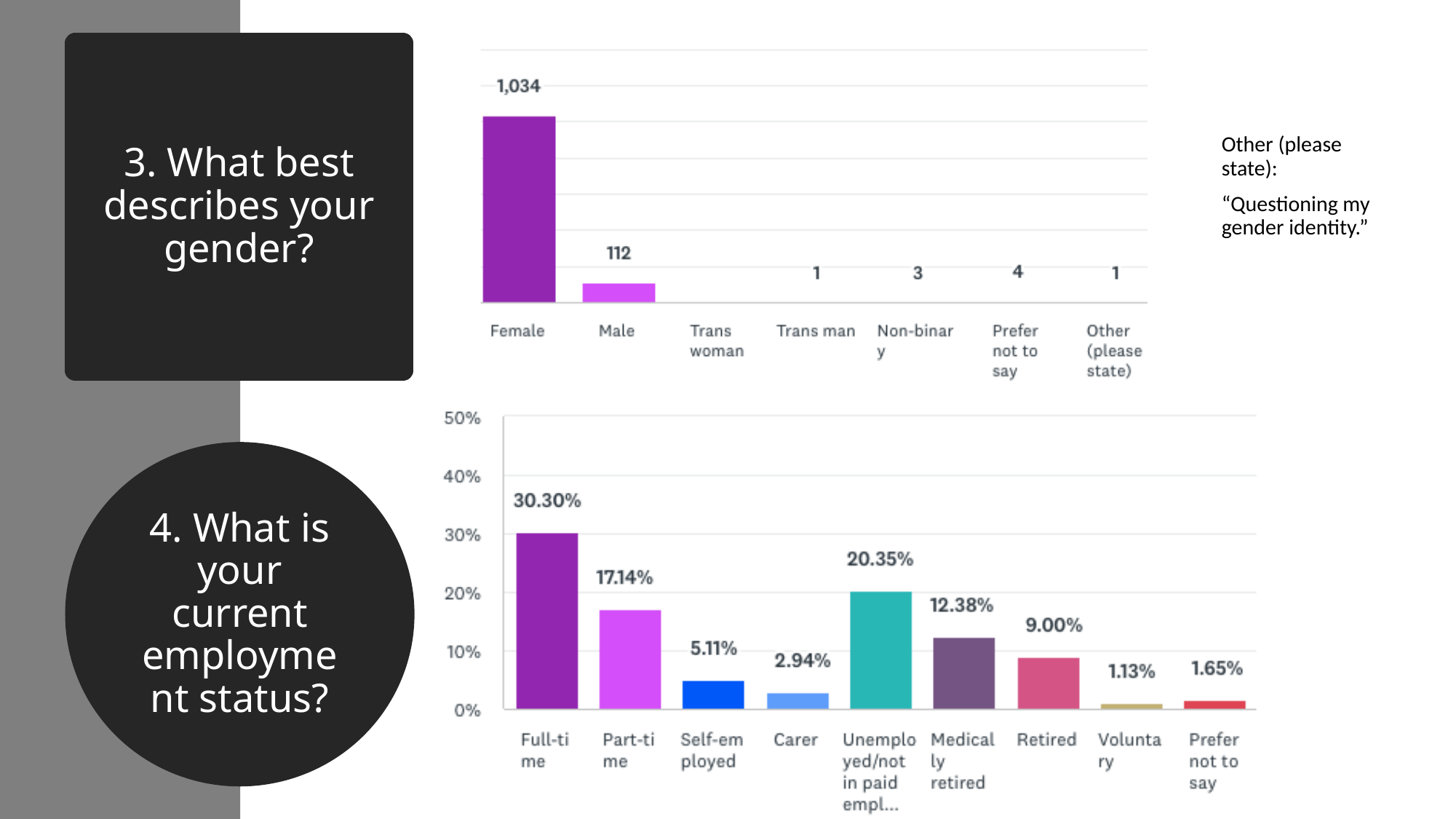

#### 3. What best describes your gender?
Other (please state):
“Questioning my gender identity.”
4. What is your current employment status?

### Slide 6
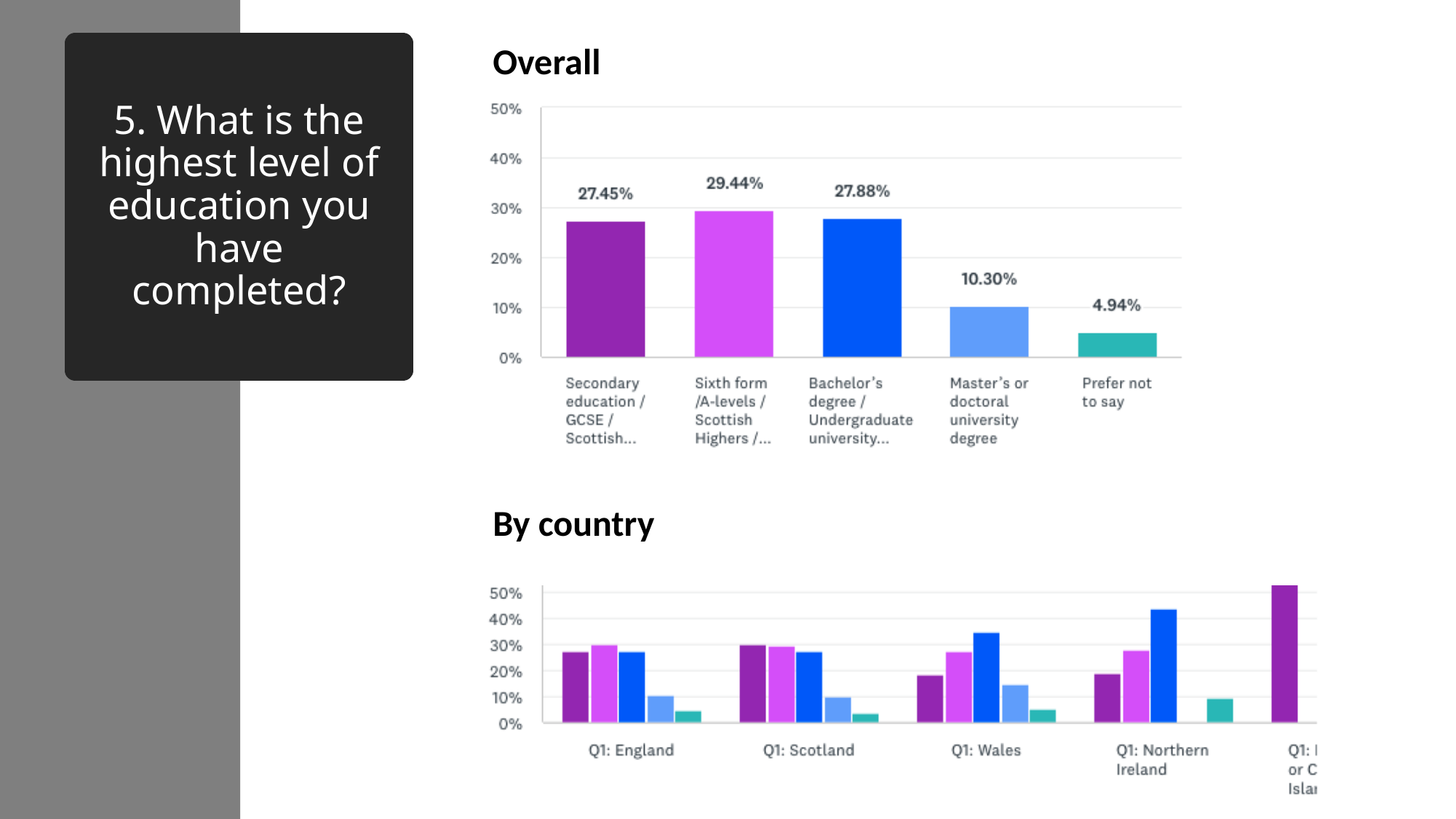

Overall
#### 5. What is the highest level of education you have completed?
By country

### Slide 7
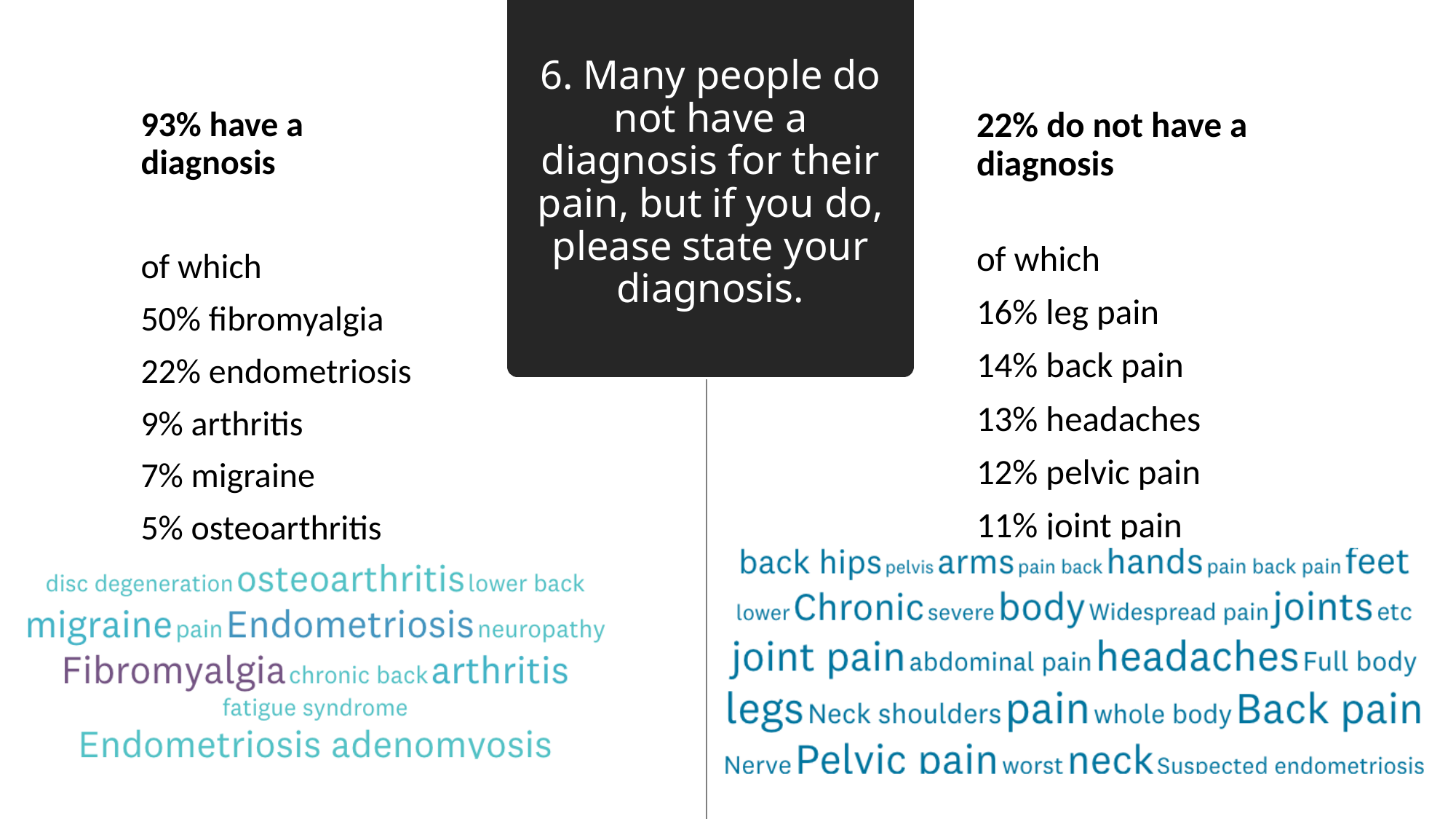

#### 6. Many people do not have a diagnosis for their pain, but if you do, please state your diagnosis.
93% have a diagnosis
of which
50% fibromyalgia
22% endometriosis
9% arthritis
7% migraine
5% osteoarthritis
22% do not have a diagnosis
of which
16% leg pain
14% back pain
13% headaches
12% pelvic pain
11% joint pain

### Slide 8
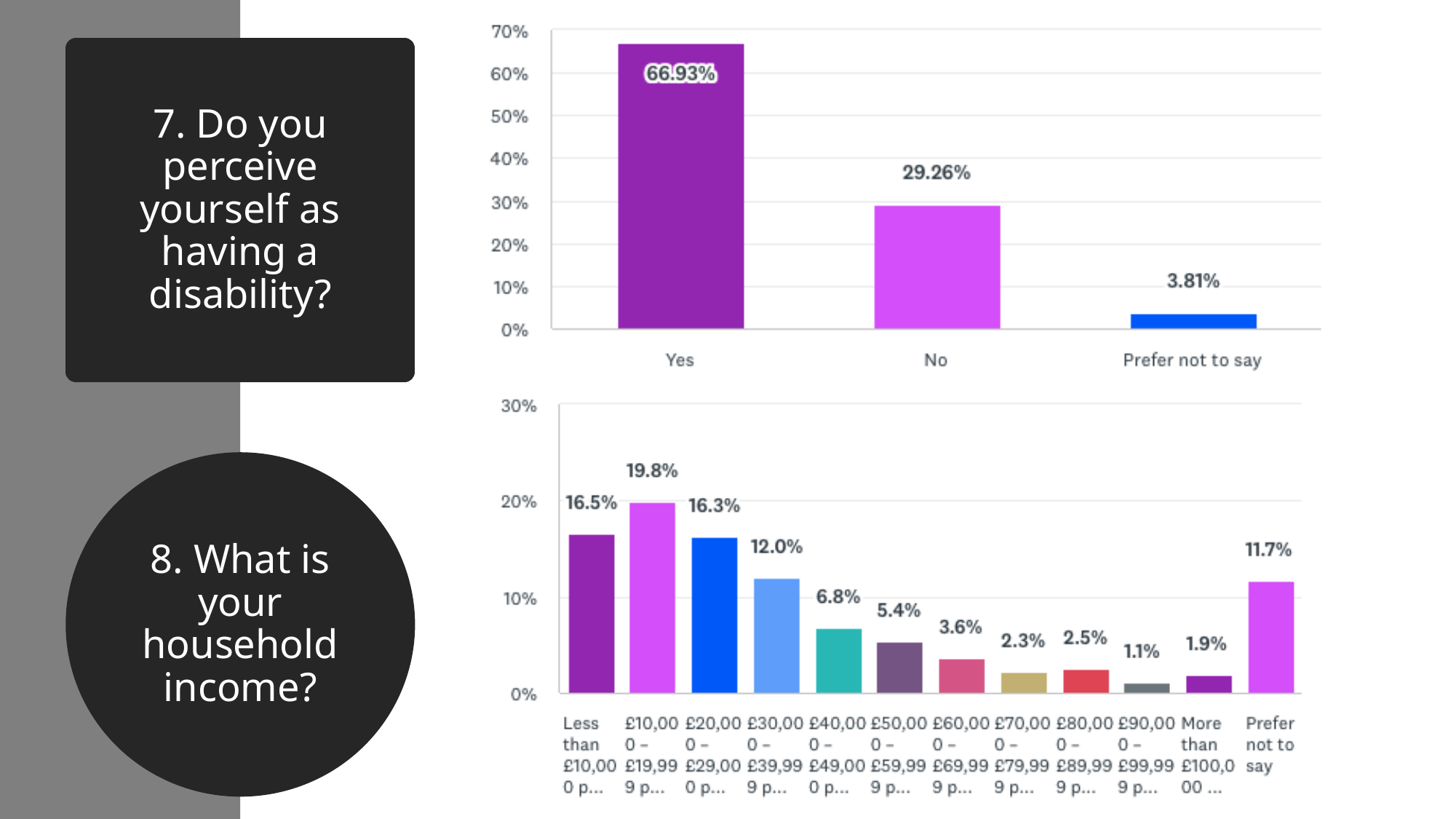

#### 7. Do you perceive yourself as having a disability?
8. What is your household income?

### Slide 9
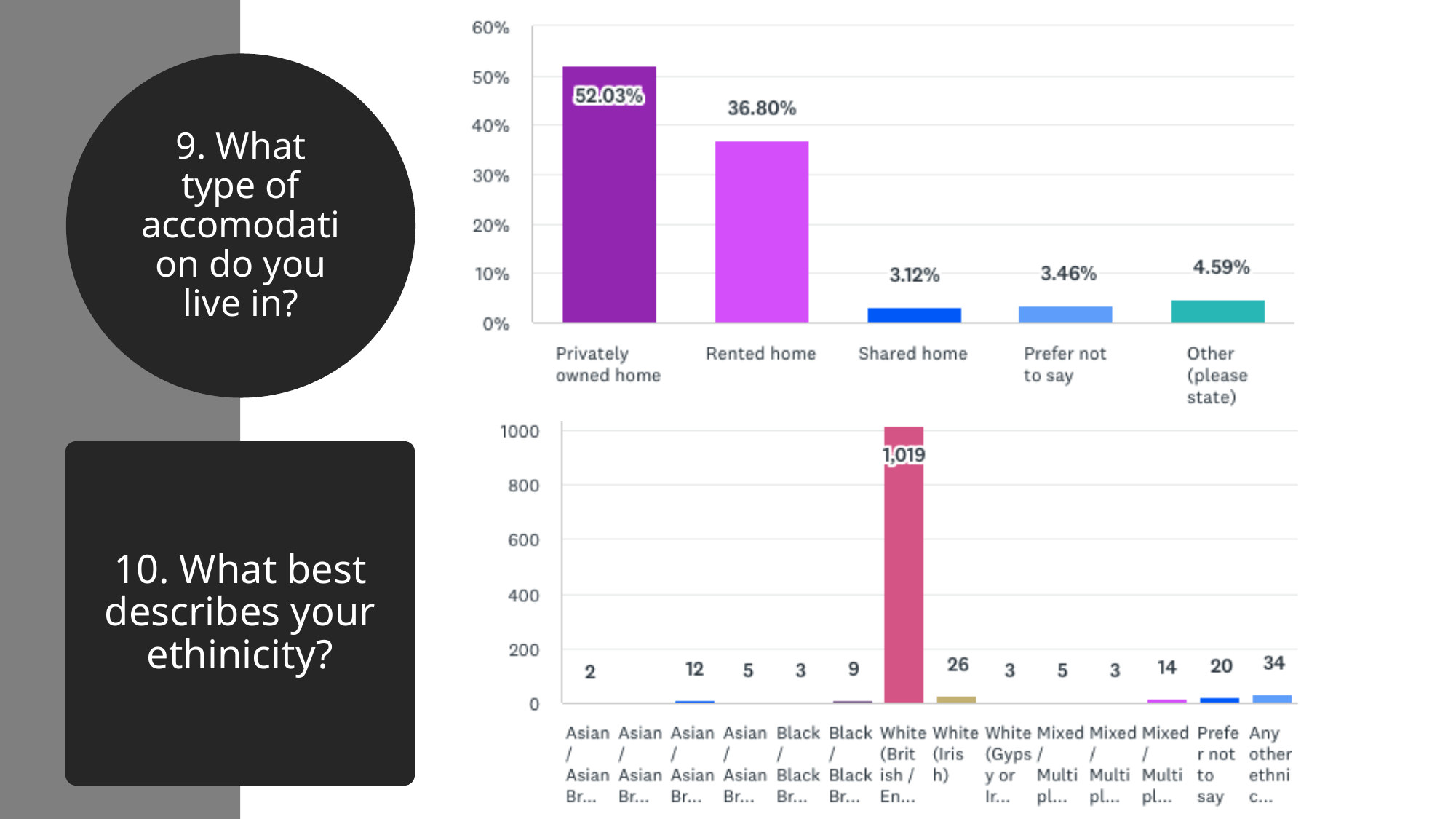

9. What type of accomodation do you live in?
#### 10. What best describes your ethinicity?

### Slide 10
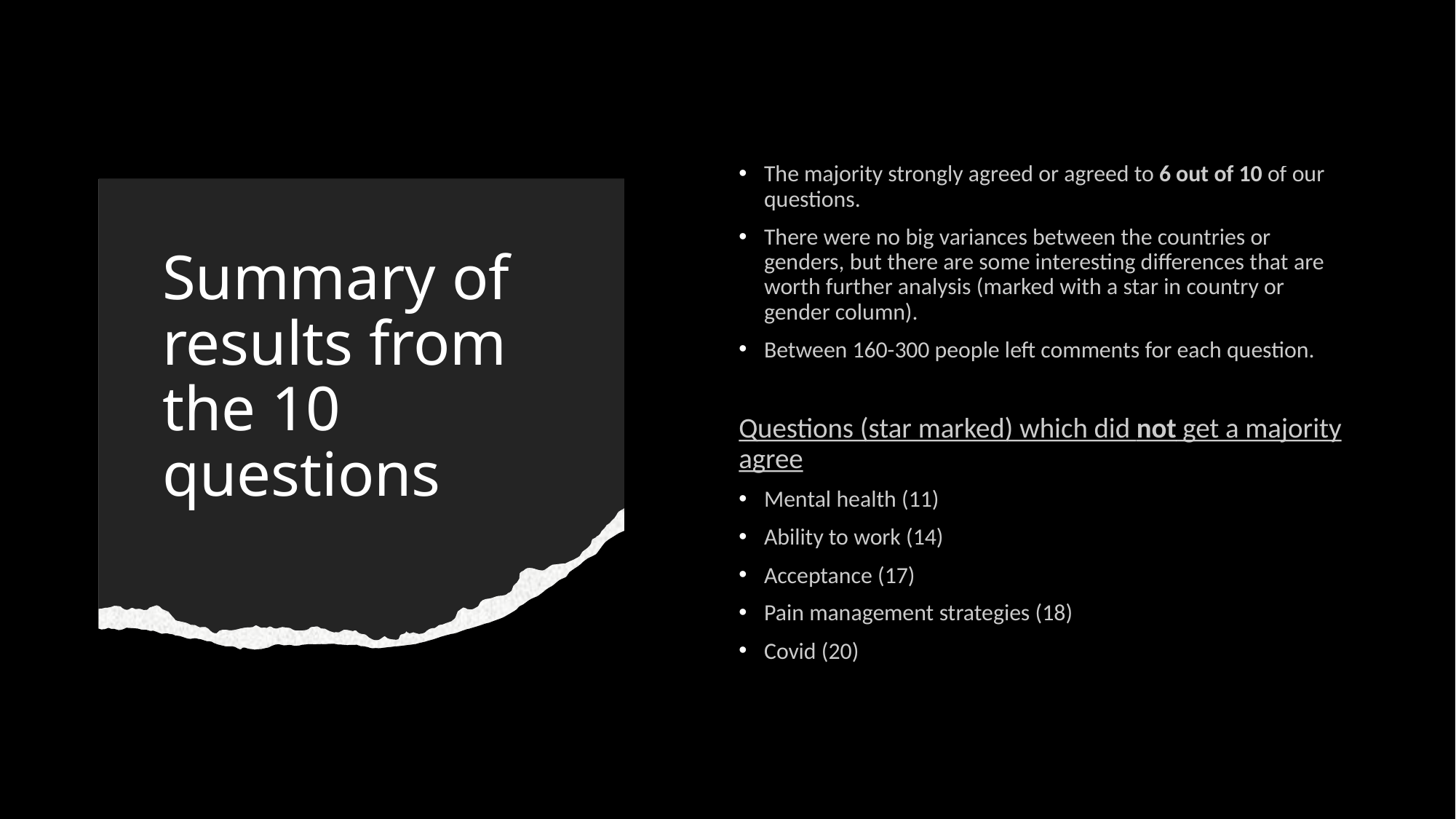

The majority strongly agreed or agreed to 6 out of 10 of our questions.
There were no big variances between the countries or genders, but there are some interesting differences that are worth further analysis (marked with a star in country or gender column).
Between 160-300 people left comments for each question.
Questions (star marked) which did not get a majority agree
Mental health (11)
Ability to work (14)
Acceptance (17)
Pain management strategies (18)
Covid (20)
#### Summary of results from the 10 questions

### Slide 11
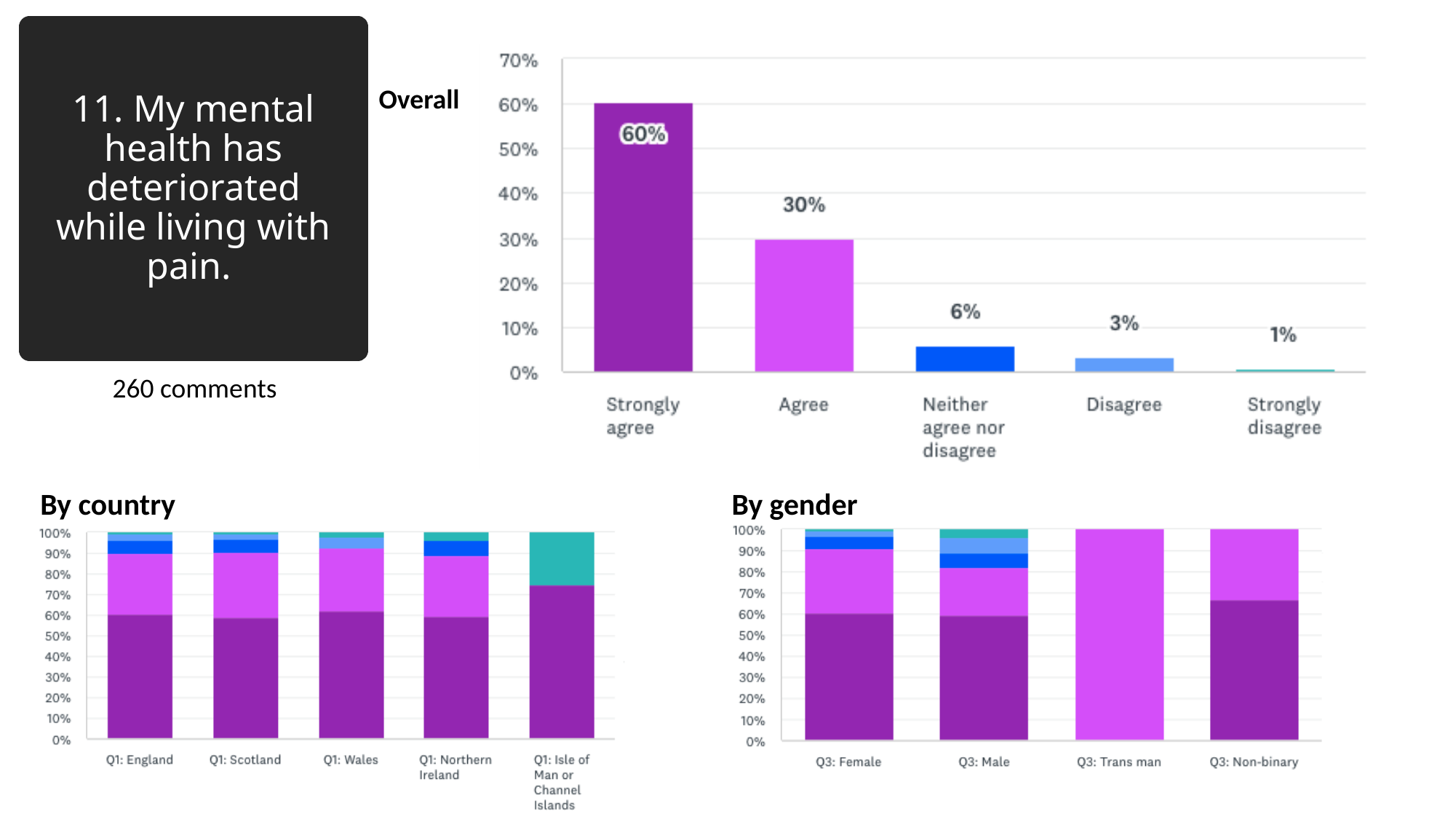

#### 11. My mental health has deteriorated while living with pain.
Overall
260 comments
By country
By gender

### Slide 12
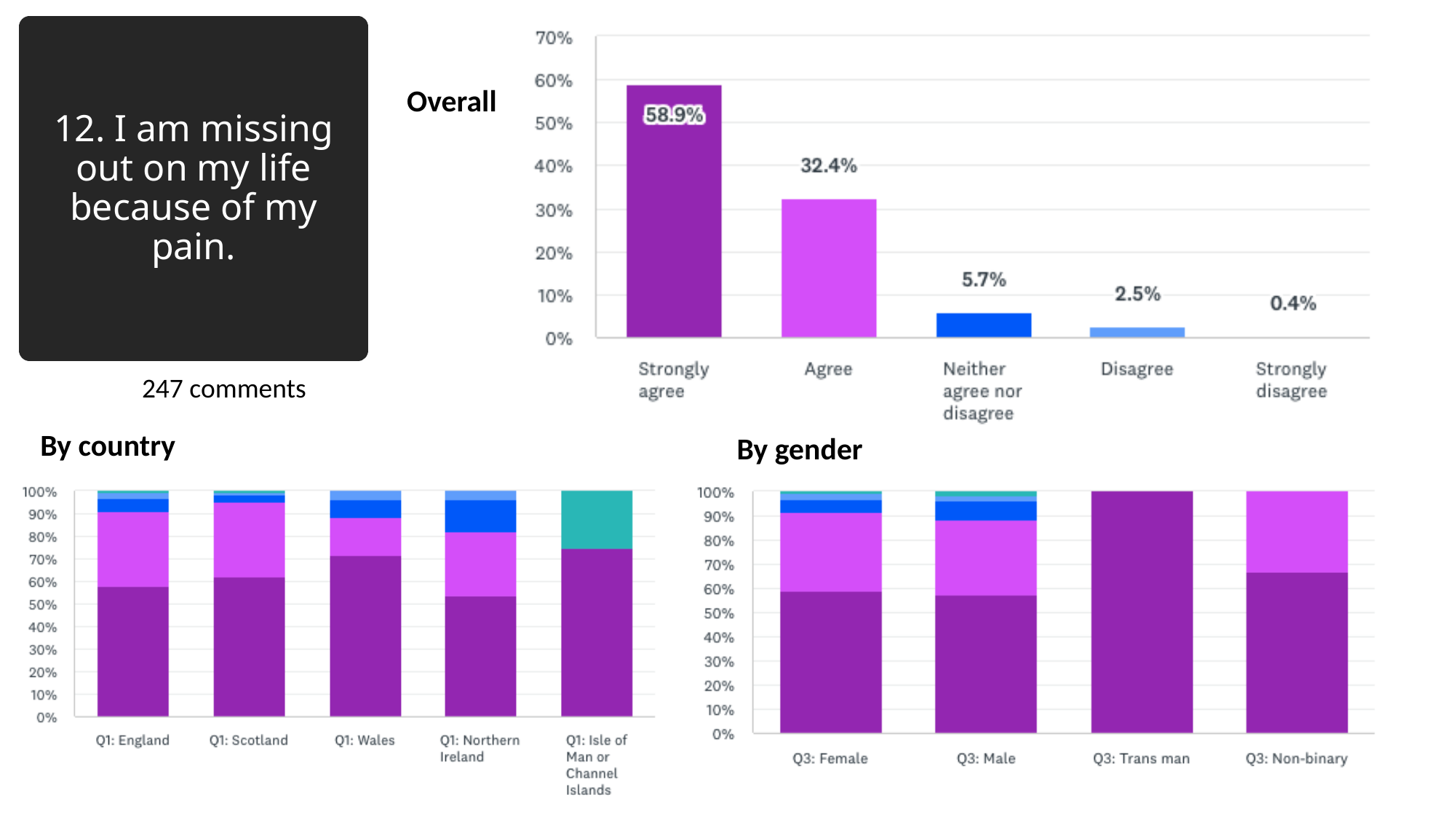

#### 12. I am missing out on my life because of my pain.
Overall
247 comments
By country
By gender

### Slide 13
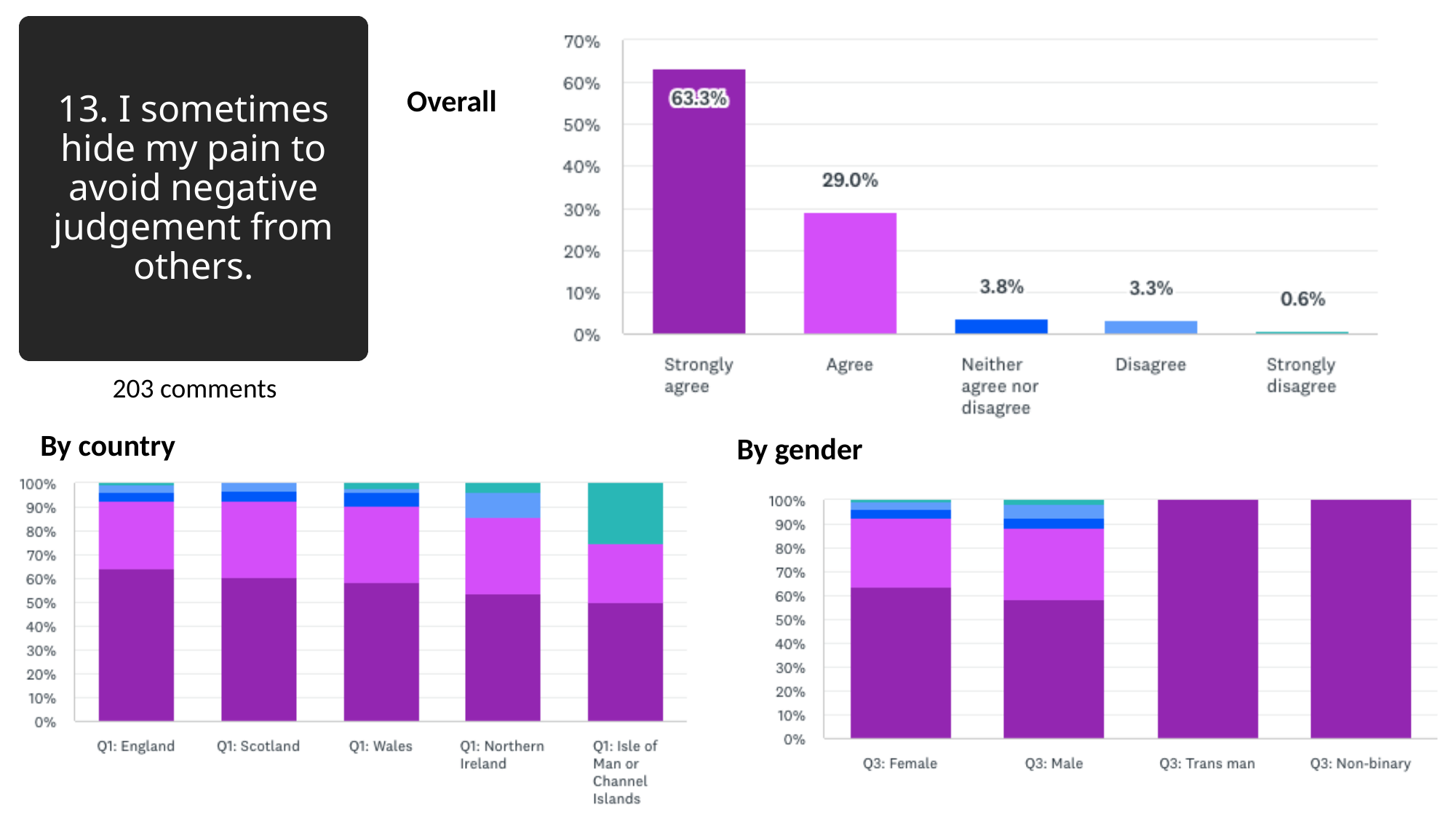

#### 13. I sometimes hide my pain to avoid negative judgement from others.
Overall
203 comments
By country
By gender

### Slide 14
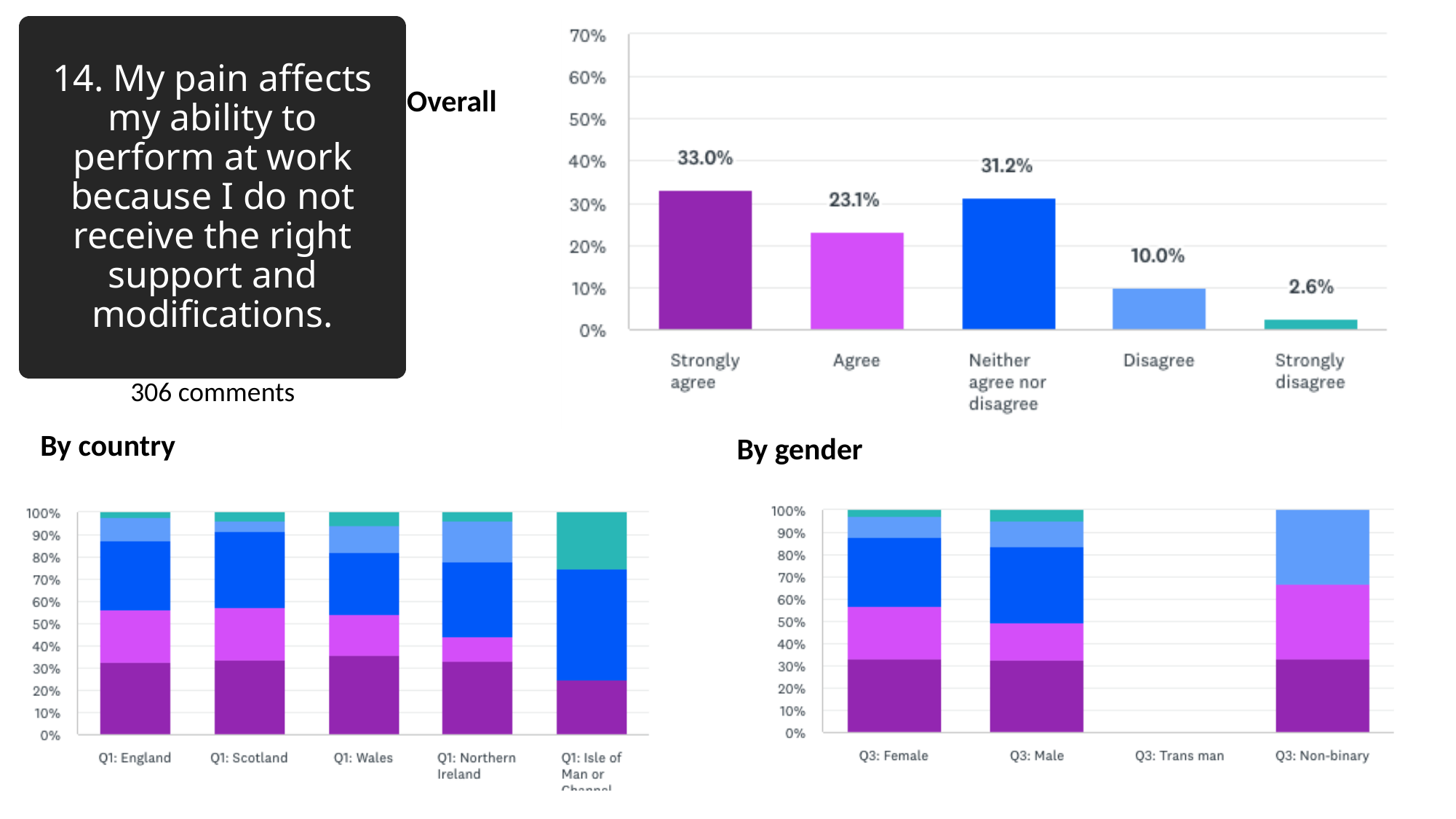

#### 14. My pain affects my ability to perform at work because I do not receive the right support and modifications.
Overall
306 comments
By country
By gender

### Slide 15
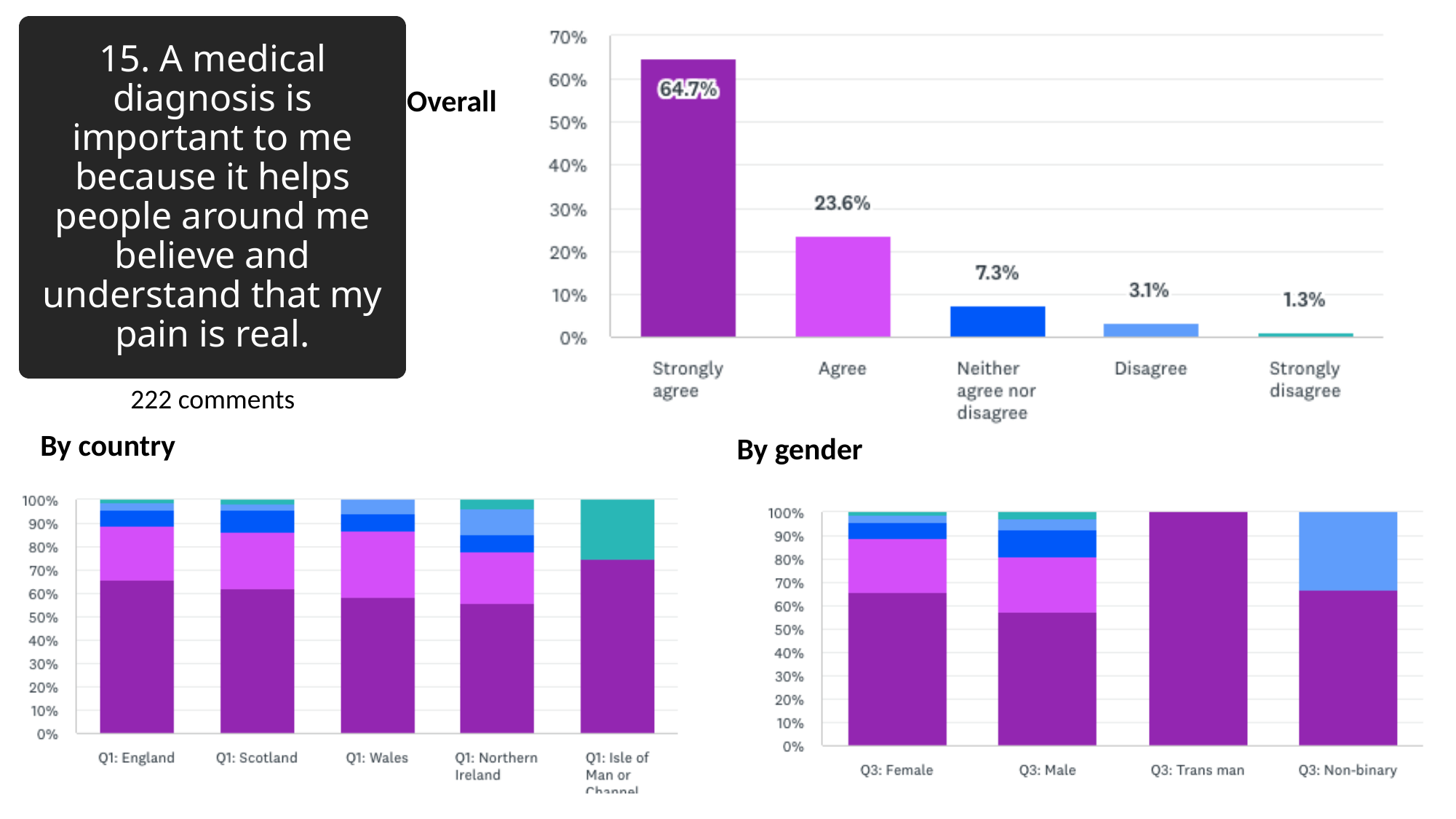

#### 15. A medical diagnosis is important to me because it helps people around me believe and understand that my pain is real.
Overall
222 comments
By country
By gender

### Slide 16
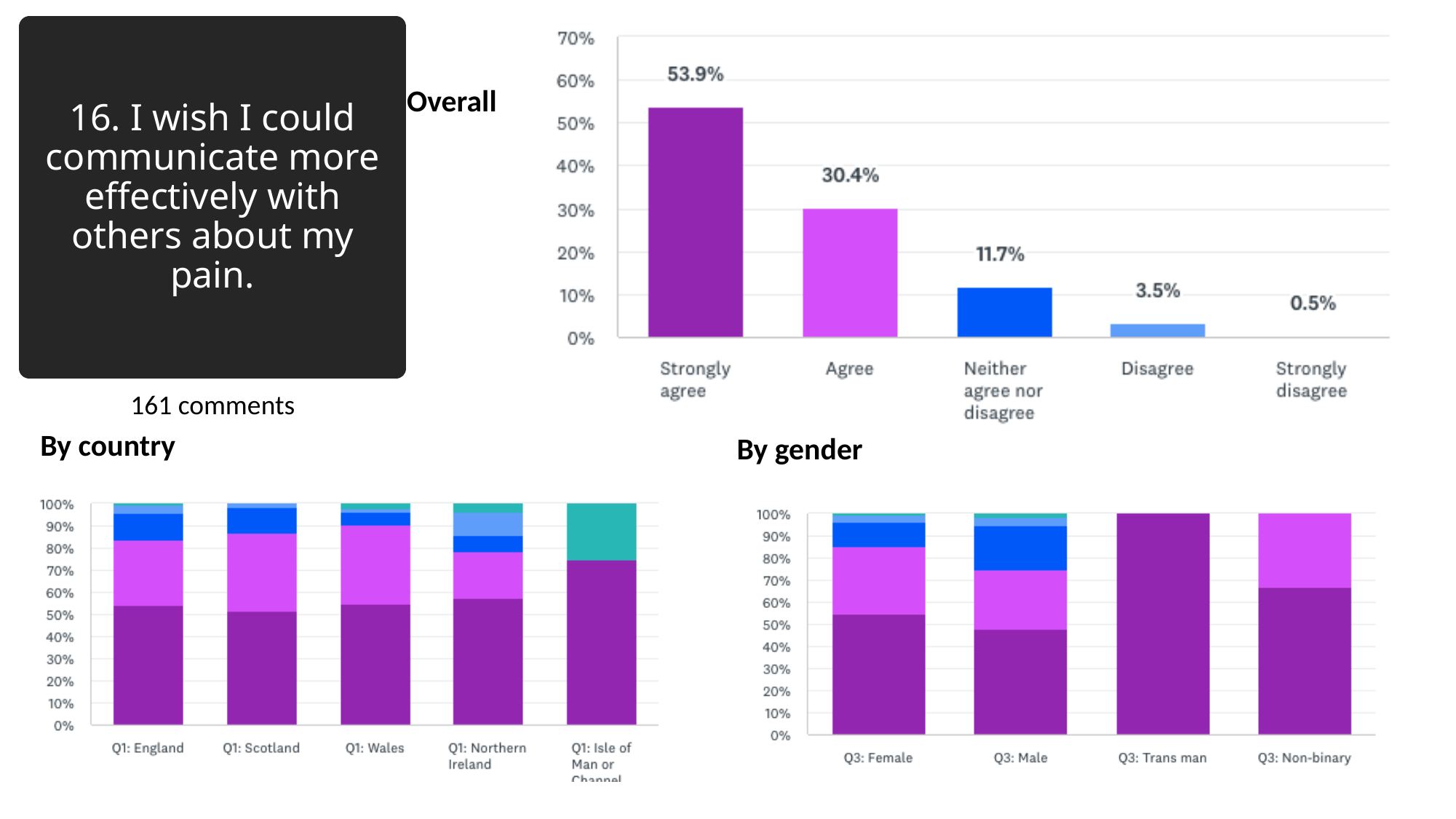

#### 16. I wish I could communicate more effectively with others about my pain.
Overall
161 comments
By country
By gender

### Slide 17
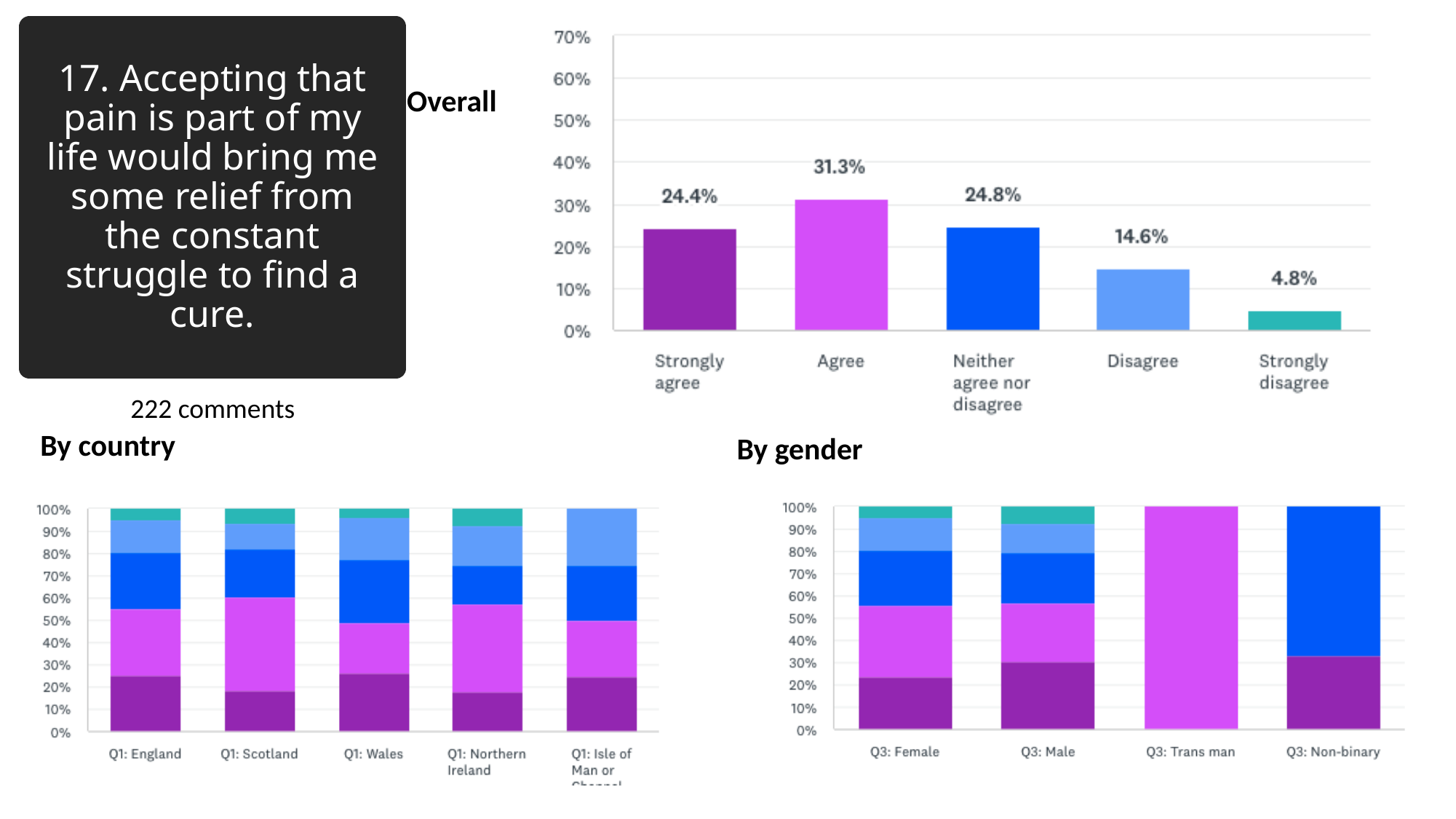

#### 17. Accepting that pain is part of my life would bring me some relief from the constant struggle to find a cure.
Overall
222 comments
By country
By gender

### Slide 18
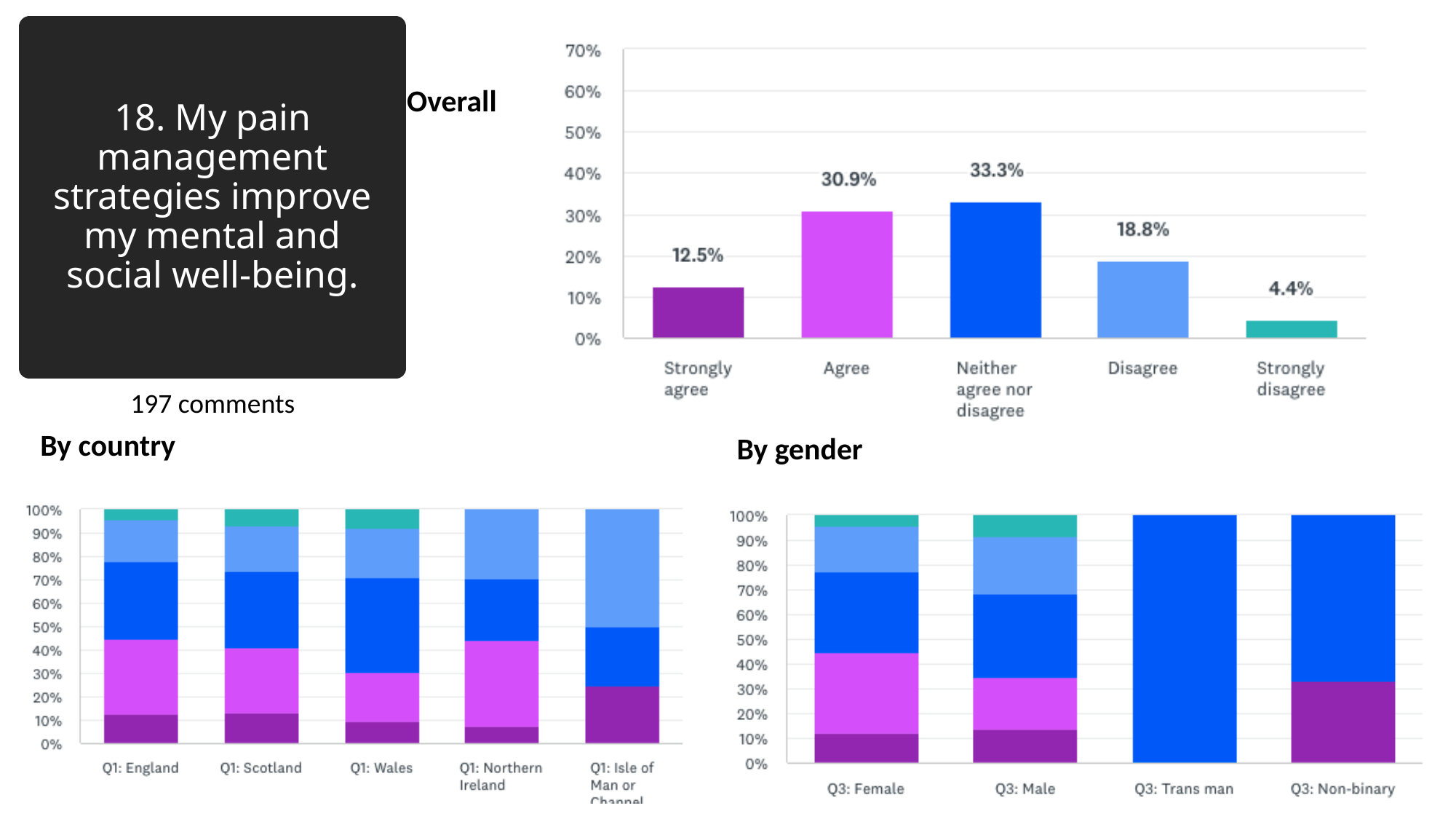

#### 18. My pain management strategies improve my mental and social well-being.
Overall
197 comments
By country
By gender

### Slide 19
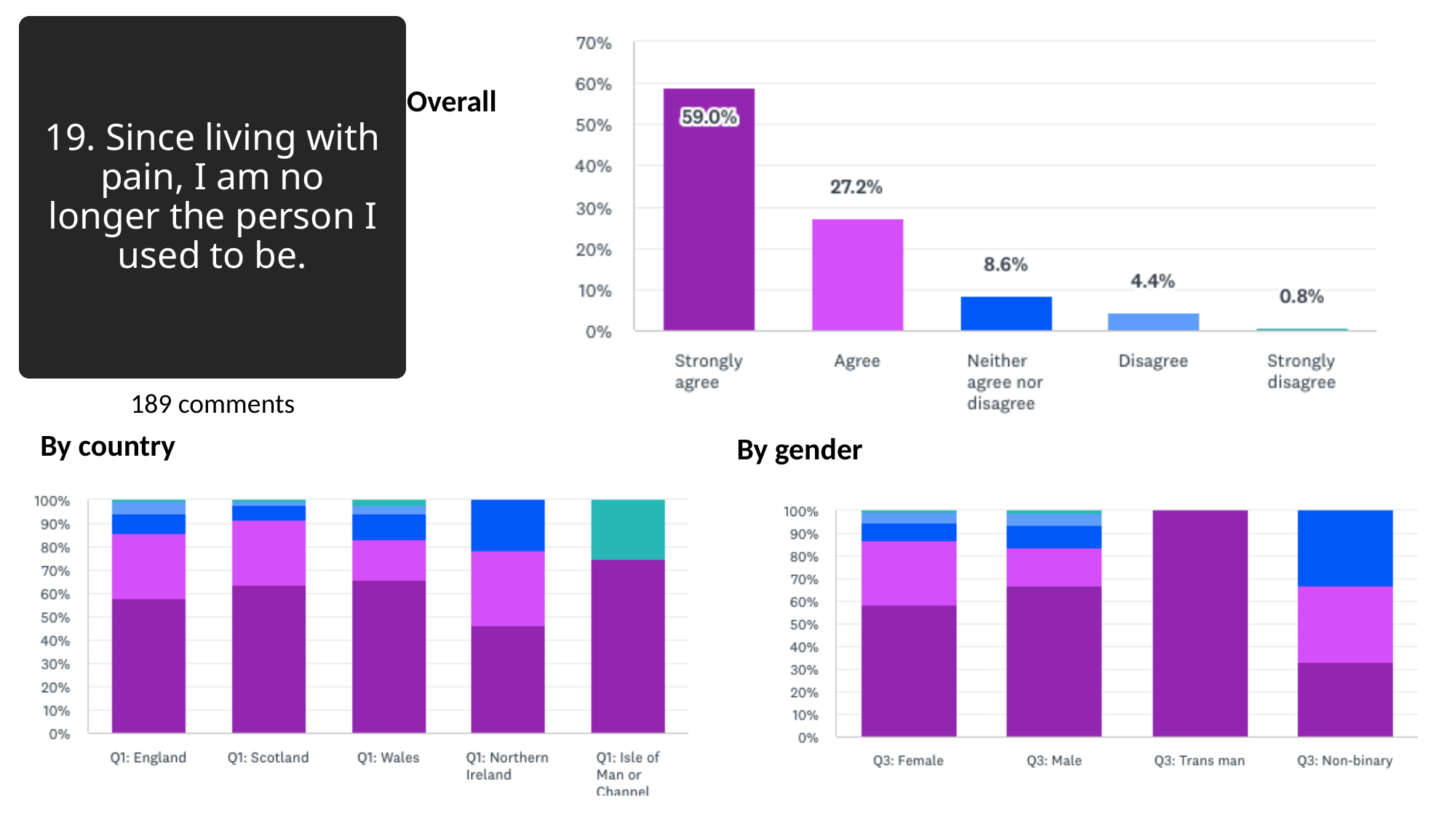

#### 19. Since living with pain, I am no longer the person I used to be.
Overall
189 comments
By country
By gender

### Slide 20
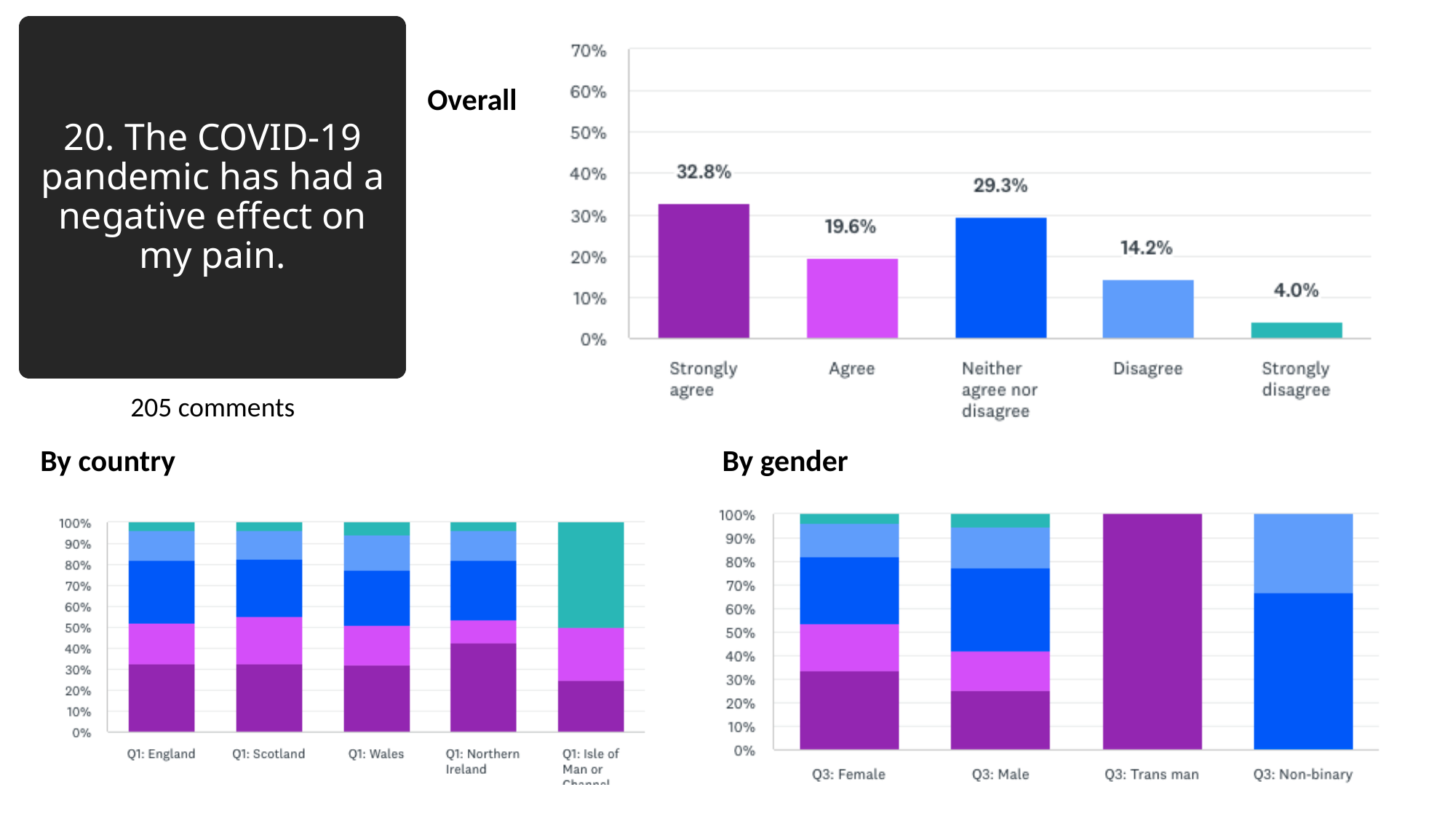

#### 20. The COVID-19 pandemic has had a negative effect on my pain.
Overall
205 comments
By country
By gender
